# Mutational signature landscape of pancreatic cancer genomes from nine countries

**DOI:** 10.64898/2026.10.21.26365999

**Authors:** Sarah Moody, Thomas Cattiaux, Jiali Pang, Haoran Zhang, Charles Nunn, Stephen Fitzgerald, Pilar Gallego-García, Priscilia Chopard, Arash Nikmanesh, Valérie Gaborieau, Christine Carreira, Calli Latimer, Saamin Cheema, Sergey Senkin, Giovana Tardin Torrezan, Anh Phuong Le, Aida Ferreiro- Iglesias, Ilinca Lungu, Marcos Díaz-Gay, Stefano Serra, Rui Manuel Reis, Denise Peixoto Guimarães, Patricia Ashton-Prolla, Alessandro Bersch Osvaldt, André de Moricz, Adhemar Monteiro Pacheco, Maria Paula Curado, Felipe José Fernandez Coimbra, Vladimir Dugalic, Simona Ognjanovic, Tatsuhiro Shibata, Satoshi Shiba, Reza Malekzadeh, Saman Nikeghbalian, Ahmadreza Niavarani, Fatemeh Shafighian, Beata Świątkowska, Janusz Strzelczyk, Katarzyna Roszkowska-Purska, Jolanta Lissowska, Ivana Holcatova, Pavel Záruba, Anush Mukeria, Oksana Shangina, David Zaridze, Samuel O Antwi, Ann L Oberg, Steven Gallinger, Julie M Wilson, Ana Carolina de Carvalho, Laura Humphreys, Sandra Perdomo, Behnoush Abedi-Ardekani, Ludmil B Alexandrov, Paul Brennan, Michael R Stratton

**Author notes:** These authors contributed equally (Sarah Moody, Thomas Cattiaux). **Corresponding Author** Correspondence to Michael R. Stratton and Paul Brennan.

## Abstract

Approximately half of all pancreatic cancers cannot be explained by known risk factors. Here we present the results of mutational signature analysis in 495 cases of pancreatic cancer from nine countries to explore whether there are previously unknown mutagenic exposures acting on pancreatic cancer genomes. The mutational signature landscape was consistent between countries. We identified one signature not previously found in pancreatic cancer, SBS111, which does not have a confirmed aetiology but is present in >99% of cases and contributes on average 26% of the total mutation burden. No evidence was found for mutational signatures known to be caused by exogenous exposures, nor was any mutational signature capable of explaining differences in incidence rates between countries. These results suggest that the unexplained causes of pancreatic cancer are unlikely to be strongly mutagenic and that non-mutagenic mechanisms may have a prominent role in explaining pancreatic cancer incidence.

## Introduction

For most human cancers the age-standardised incidence rates (ASR) show geographic variation, with these trends often changing over time. In many cases the factors driving these differences are unknown. The most likely explanation is that these are the result of environmental or lifestyle factors which influence the risk of developing cancer and which are also, in theory, preventable^1^. Whilst traditional cancer epidemiology has successfully identified many such cancer risk factors, new discoveries have slowed over time and new approaches are needed to complement this strategy. One such approach is the concept of mutational epidemiology, which combines cancer epidemiology with the field of cancer genomics. Mutational signatures are the consequence of mutational processes imprinting specific patterns of mutations across the genome. Whilst some mutational processes are the result of endogenous cellular functions such as those linked to DNA replication and repair, others are caused by exogenous mutagenic exposures such as UV radiation and tobacco smoke^2^. Previous studies of large numbers of cancer genomes have established a reference set of mutational signatures but the aetiology for many remains unknown^2^. By analysing large numbers of cancer genomes across countries with varied incidence rates, it is possible to identify signatures which are geographically restricted and/or associate with the incidence rate. Such signatures likely represent exogenous exposures which influence the risk of cancer in exposed individuals. Utilising this type of experimental design, the Mutographs Cancer Grand Challenge project has yielded insights into mutagenic exposures present in other cancer types, including kidney and colorectal cancers^3–7^.

The incidence rate of pancreatic cancer varies between countries, with the highest rates of 8-10 per 100,000 being typically observed in Central and Northern Europe, as well as Japan and North America. These rates are around double what is observed in most middle-income countries. Incidence rates have also been increasing gradually over the past two decades in some but not all countries, with the most rapid increases occurring in parts of Asia and Europe^8–11^. These increases are observed in both early onset cases (individuals under the age of 50) and in older individuals^12,13^. The known modifiable risk factors for pancreatic cancer include tobacco, alcohol, obesity, diabetes and pancreatitis, although together these only account for approximately one third of the total disease burden^12,14–16^. Inherited risk factors also play an important role^15^, but even when taking these into account a substantial proportion of pancreatic cancers are unexplained suggesting the presence of additional, previously undetected, environmental or lifestyle risk factors^16^.

Previous whole-genome sequencing studies conducted on pancreatic cancers did not have the ability to examine geographic variation in the mutational signature landscape due to the cohorts being limited to a single country or region^17,18^. Here, we present the results of mutational signature analysis on 495 cases of pancreatic cancer from nine countries, to investigate whether there are previously unknown mutagenic exposures acting on pancreatic cancer genomes and whether variation in mutational processes contributes to pancreatic cancer incidence rates.

## Results

### Data collection

A total of 533 cases of pancreatic cancer were collected from 9 countries across 4 continents, including from Canada (n = 171), United States (n = 71), Russia (n = 60), Czech Republic (n = 60), Poland (n = 50), Iran (n = 50), Japan (n = 30), Serbia (n = 22) and Brazil (n = 19) (**Fig. 1, Supplementary Table1**). In most cases, laser capture microdissection was used to enrich tumour cell content (**Methods**). Whole genome sequencing was performed on tumour and matched normal samples with median coverage of 53x and 27x respectively. In line with previous Mutographs studies, cases with canonical DNA repair deficiencies or evidence of signatures from previous chemotherapy treatment were excluded from the analysis to avoid interference with signature extraction (**Supplementary Note**)^4^. In addition, a single hypermutated case with a distinct mutational spectrum was excluded (**Supplementary Note**). A total of 495 cases were retained for further analysis.

**Fig. 1.**
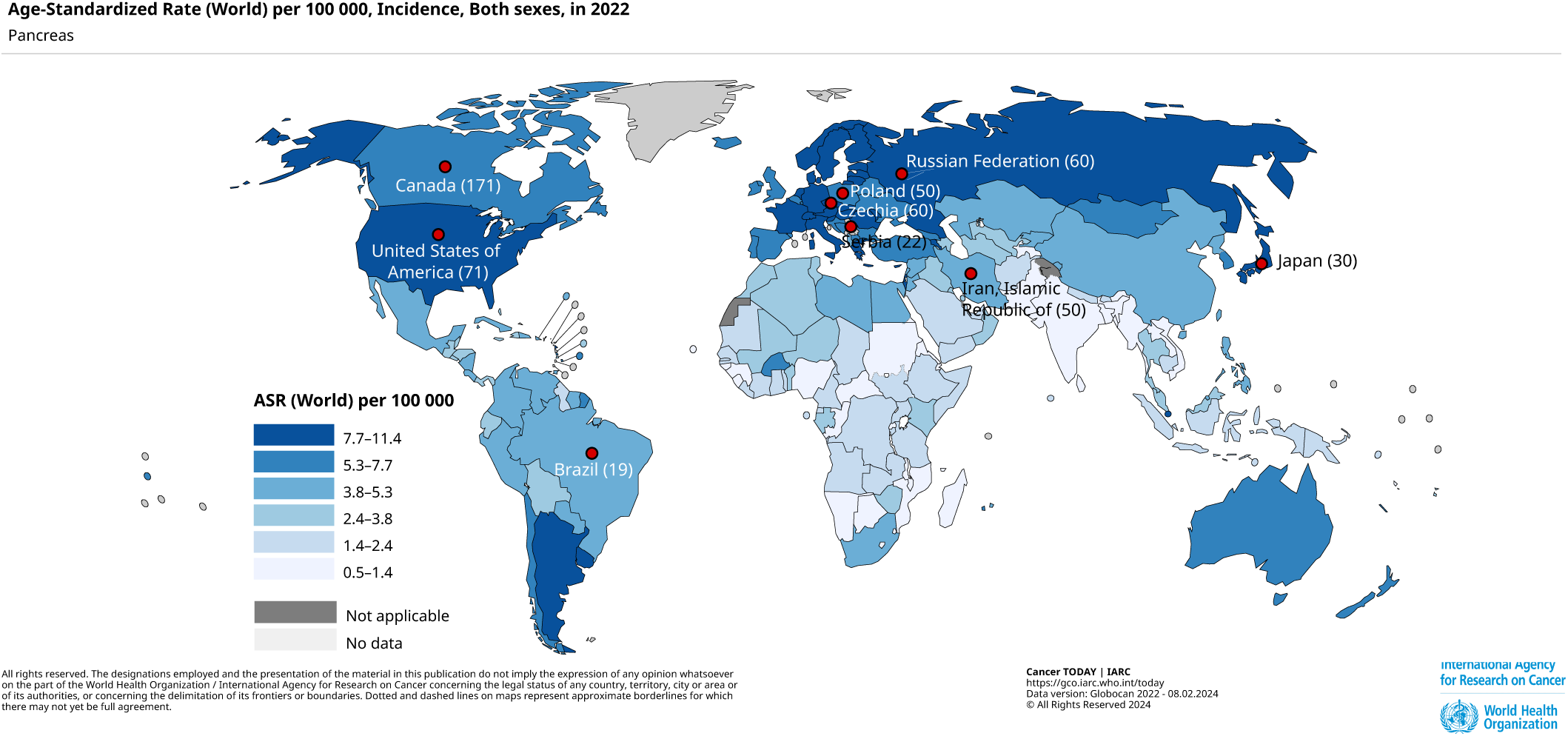
Pancreatic cancer incidence rates and data collection. Incidence of pancreatic cancer for men and women combined (ASR per 100,000). Data from GLOBOCAN 2022. Markers indicate countries included in this study (number of participants per country).

### Mutation burdens

The somatic mutation burdens for the 495 pancreatic cancers ranged from 1,051 to 30,933 (median 4,216) for single base substitutions (SBS) and from 60 to 3,365 (median 580) for small insertions and deletions (indels or ID) (**Supplementary Table 2**). No significant differences were observed between countries after corrections for tumour purity (**Extended Data Fig. 1a**). The most common driver mutations were *KRAS* (92%) and *TP53* (78%) as previously reported (**Supplementary Table 3, Supplementary Note**). No significant differences in overall driver frequency or specific *KRAS* variants were observed between countries (**Extended Data Fig. 1b, Supplementary Note**). Principal component analysis was conducted on the relative proportions of the SBS96 mutation classes which showed no clear separation of any individual country (**Supplementary Table 4, Extended Data Fig. 1c**).

### Mutational signature landscape

Mutational signature analysis was performed using two methods, SigProfilerExtractor and mSigHdp^19,20^ (**Fig. 2, Supplementary Tables 4 & 5, Supplementary Note**). The extracted *de novo* mutational signatures were then decomposed into known reference signatures in the Catalogue of Somatic Mutations in Cancer (COSMIC) database version 3.6 (**Supplementary Table 6, Supplementary Note**). We identified 12 SBS mutational signatures, all of which were composed of reference signatures in the COSMIC database, including SBS1 (spontaneous deamination of 5-methylcytosine); SBS2 and SBS13 (Apolipoprotein B mRNA editing enzyme catalytic polypeptide-like [APOBEC] family activity); SBS5 (unknown age-related); SBS8 (unknown); SBS17a and SBS17b (unknown); SBS18 (reactive oxygen species), SBS40a (unknown, age-related), SBS93 (unknown) and SBS103 (unknown)(**Fig. 2a**)^2,21^. The last signature was SBS111 which was recently reported in microsatellite stable colorectal cancers and is thought to be linked to DNA repair infidelity^4,22^. We also identified 10 indel signatures from the COSMIC reference database (**Supplementary Table 7, Supplementary Note)**, including ID1 and ID2 (polymerase slippage on the nascent or template strand respectively); ID8 and ID17 (speculated to be related to repair of double stranded breaks in DNA and/or mutations in TOP2A) and ID4, ID5, ID9, ID12, ID14 and ID21 (all of unknown aetiology, **Fig. 2b**)^2,21^.

**Fig. 2.**
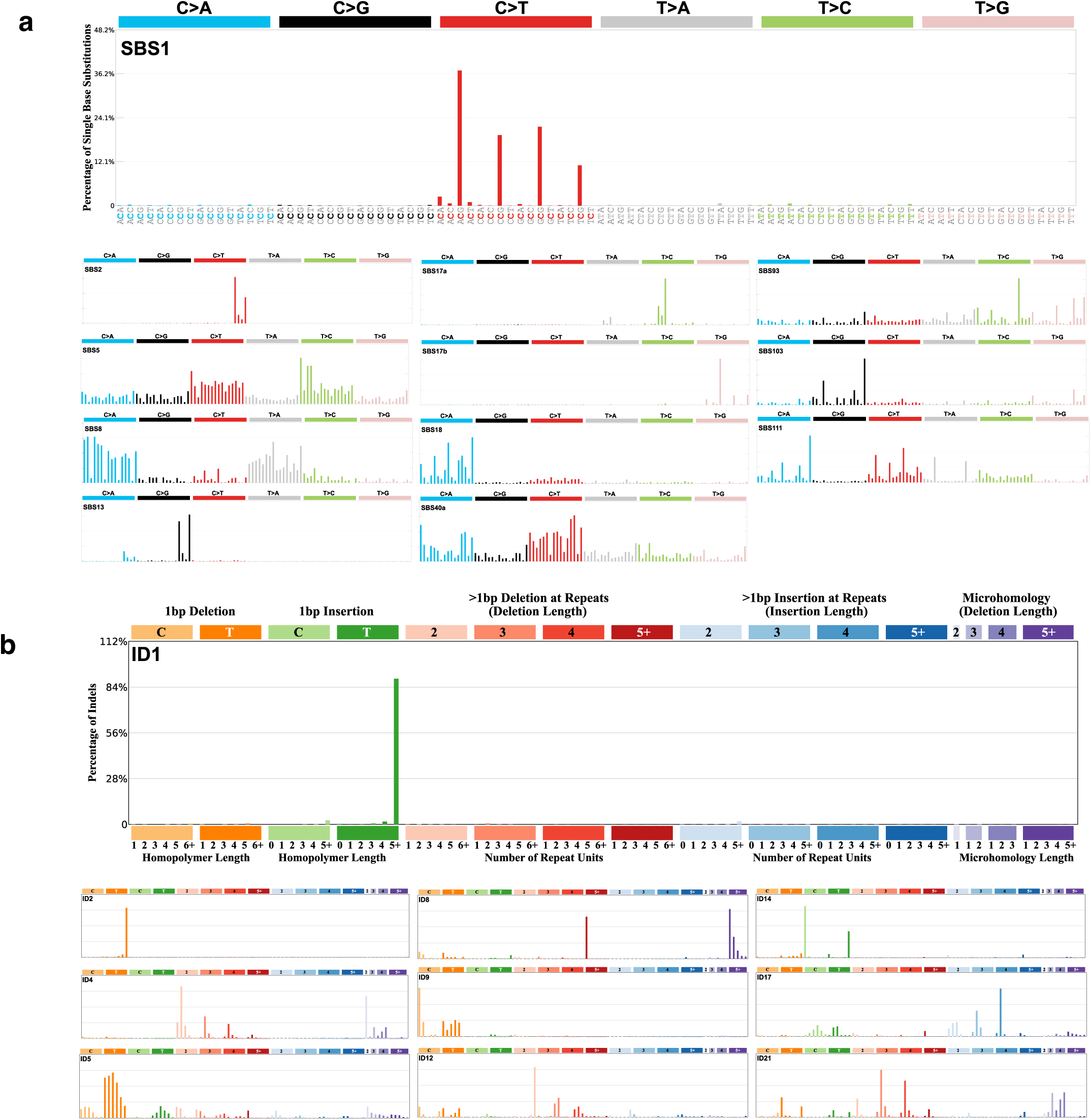
Mutational signatures active in pancreatic cancer genomes. Mutational signatures extracted from 495 cases of pancreatic cancer genomes including a) single-base substitution signatures present in the Catalogue of Somatic Mutations in Cancer (COSMIC) database of reference mutational signatures and b) indel signatures present in the Catalogue of Somatic Mutations in Cancer (COSMIC) database of reference mutational signatures

### Burden of mutational signatures

The activity of each signature in each sample was assigned using MSA (**Supplementary Tables 8-9**)^23^. Four mutational signatures were present in the majority of cases and contributed over 80% of the average total SBS mutation burden, SBS111 (493/495 cases, 26%), SBS40a (485/495 cases, 26%), SBS1 (494/495 cases, 19%) and SBS5 (333/495 cases, 11%)(**Fig. 3a**). There was no significant enrichment of any SBS signature in any individual country (**Fig. 3b&e, Supplementary Table 10**). For indel signatures ID1 and ID2 made up 61% and 22% of the average total indel burden (**Fig. 3c&d**). Again, all signatures were consistent across countries except for ID12. This signature was rare in Japanese cases (1/29 cases, 3% - OR = 0.014, q-value = 1.1 × 10−7) despite being common in those from elsewhere (403/466 cases, 86%), in addition to being enriched in US cases (OR = 8.689, q-value = 1.4 × 10−6). (**Fig. 3e, Supplementary Table 10**). As was previously observed in colorectal cancers, SBS111 was significantly correlated with both total Indel burden and ID1 burden (**Extended Data Fig. 2**)^22^.

**Fig. 3.**
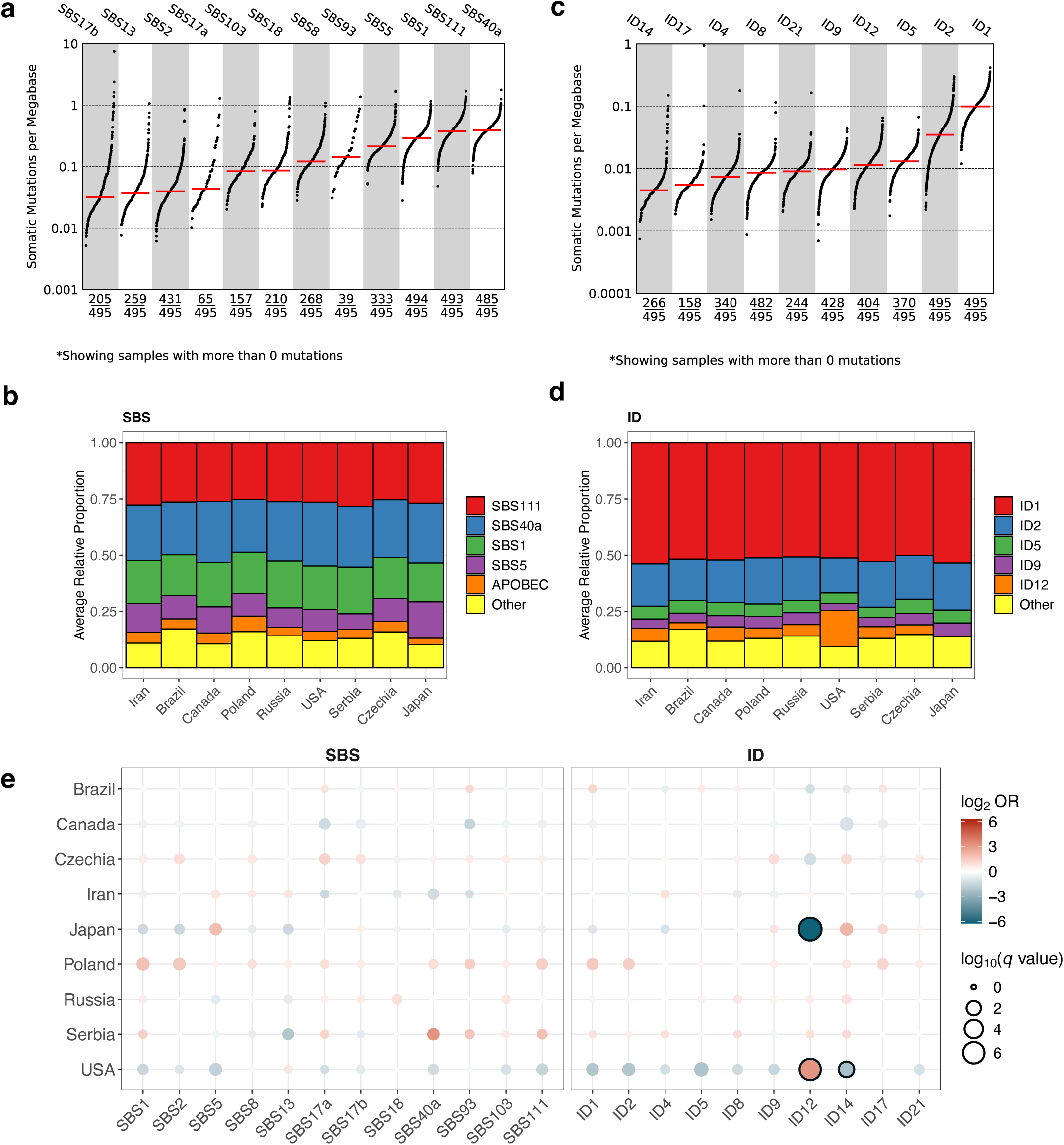
Burden of mutational signatures in pancreatic cancer genomes. **a)** TMB plot showing the number of mutations per megabase and frequency for each SBS mutational signature. Data include only samples with more than zero mutations. **b)** average mutation burden of SBS mutational signatures in each country. All individual mutational signatures that contribute less than 5% on average are grouped in the ‘others’ category. **c)** TMB plot showing the number of mutations per megabase and frequency for each Indel mutational signature. Data include only samples with more than zero mutations. **d)** average mutation burden of ID mutational signatures in each country. All individual mutational signatures that contribute less than 5% on average are grouped in the ‘others’ category. **e)** Variation in signature across countries relative to all others. Multivariate logistic regression models adjusted for age at diagnosis, sex, and tumour purity were used to test for enrichment. Firth’s bias-reduced logistic regression was applied when complete or quasi-complete separation occurred or when standard models did not converge. Points are coloured by odds ratio (OR), with point size indicating statistical significance. P values were corrected using the Benjamini– Hochberg procedure and reported as q values; tests with q < 0.01 are outlined in black.

Compared to previous studies, SBS5 made up a smaller proportion of the average total SBS mutation burden in our cohort^2^. This is most likely due to SBS111 not being extracted previously and thus the mutations that would have been assigned to it were instead assigned to SBS5. However, given that both signatures generate a broad pattern of mutations across many contexts it is possible that technical artefacts cause SBS5 mutations to be inappropriately assigned to SBS111.

### Validation in simulated datasets and external cohorts

In order to validate our findings, we used simulations and external datasets. To provide a known ground truth from which it is possible to establish how accurately the attribution algorithm is able to distinguish between SBS5 and SBS111, we generated a simulated version of the pancreatic cancer dataset and attributed the SBS signature panel using MSA (Methods). This showed that the average attributed levels of SBS5 and SBS111 matched the ground truth (**Extended Data Fig. 3a&b, Supplementary Table 11**). We also conducted a false positive test where SBS111 was omitted from the simulated dataset but included in the attributed panel of signatures. SBS111 was not attributed to any cases in this scenario (**Supplementary Table 12**). Finally, we also confirmed that SBS111 can be extracted from 318 cases with publicly available data from the ICGC, Hartwig and GEL pancreatic cancer cohorts (**Extended Data Fig. 3c**)^24^. These findings support the robustness and reproducibility of SBS111 attribution and indicate that SBS111 can be distinguished from SBS5 under the conditions tested.

### Evolutionary analysis

Mutational processes that contribute to the early stages of cancer development should be present in the early clonal mutations of cancer clones. In order to investigate the timing of mutational processes in pancreatic cancer we performed attributions independently to early clonal and late clonal mutations (**Methods**). This showed that the endogenous signatures SBS5, SBS40a and SBS1 were the most enriched in early clonal mutations (**Fig. 4a, Supplementary Table 13-17**). SBS111 was enriched in late clonal mutations as previously reported in colorectal cancers (**Fig. 4a&b**). Despite this enrichment and in contrast to colorectal cancers, SBS111 was also active in the early clonal mutations of almost all pancreatic cancers (180/189, 95%, **Fig. 4c**)^22^.

**Fig. 4.**
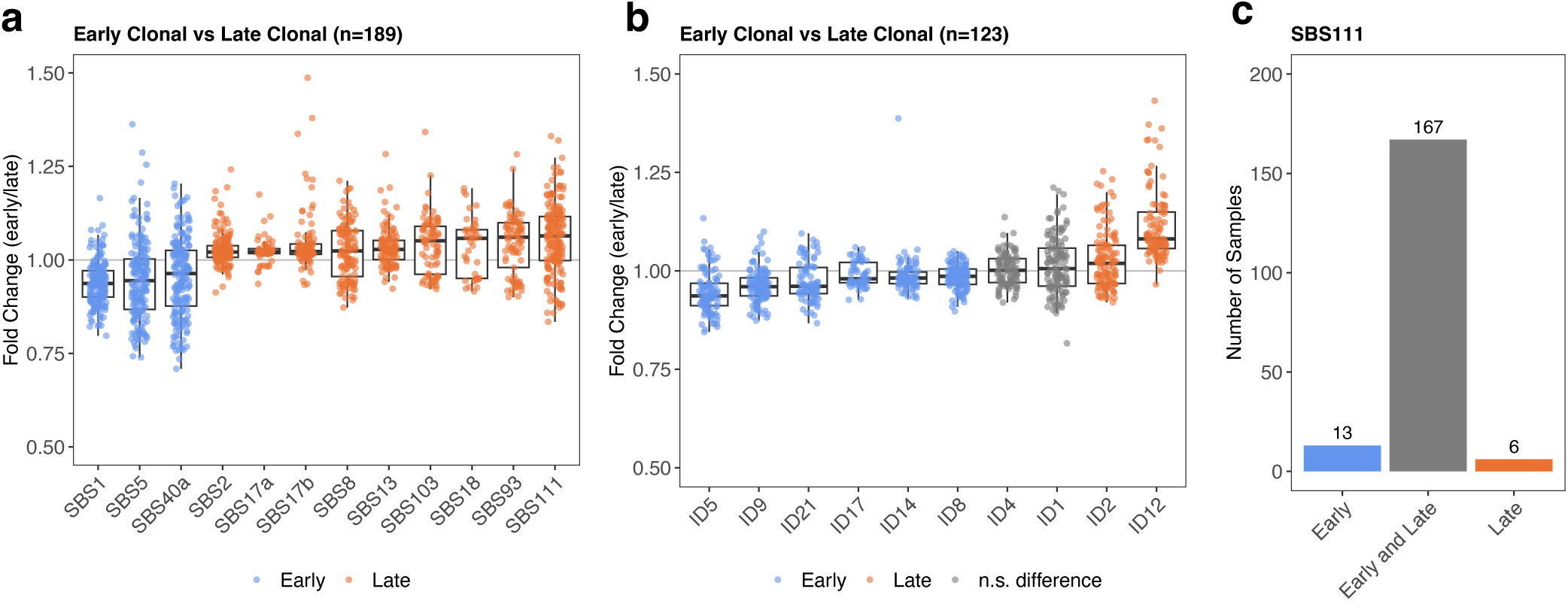
Timing of mutational signatures in pancreatic cancer genomes. Timing of **a)** SBS and **b)** ID mutational signatures showing the fold change in relative contribution between early clonal and late clonal mutations. Signatures are sorted by median fold change and coloured according to whether there is significant enrichment in early/late clonal mutations or no significant enrichment. Significance was calculated using a two-sided Wilcoxon signed-rank test with resulting p values corrected using the Benjamini-Hochberg procedure and reported as q values. Box-and-whisker plots are in the style of Tukey. The line within the box is plotted at the median, while upper and lower ends indicate 25th and 75th percentiles. Whiskers show 1.5 × IQR. **c)** Number of cases positive for SBS111 in early clonal only, both early and late clonal and late clonal only. Cases negative in both early and late are omitted.

### Associations with epidemiological data

No mutational signatures demonstrated a statistically significant association with ASR capable of explaining the observed differences in incidence (**Fig. 5a, Supplementary Table 18, Supplementary Note**). Similarly, there were no significant associations between COSMIC signature attributions and any individual risk factor including tobacco or alcohol consumption, BMI, or history of diabetes or pancreatitis, after adjustment for age, sex, country, and tumour purity (**Fig. 5b, Supplementary Table 19**). A significant association was seen between SBS5, ID5, ID8 and ID9 with age of diagnosis (**Fig.5b**), consistent with previous reports of the clock-like behaviour of these signatures^2^. No significant associations were seen between any mutational signature and genetic ancestry apart from the depletion of ID12 in individuals of East Asian ancestry, which is due to the signature being largely absent in Japan (**Fig. 5c, Supplementary Table 20**). No significant enrichment of any mutational signatures was observed in early-onset pancreatic cancers compared to those of later onset (**Fig. 5d, Supplementary Table 21)**. While this analysis was sufficiently powered to detect moderate to large effects, it is not possible to rule out the presence of modest effects below the threshold determined by power calculations (**Supplementary Note**).

**Fig. 5.**
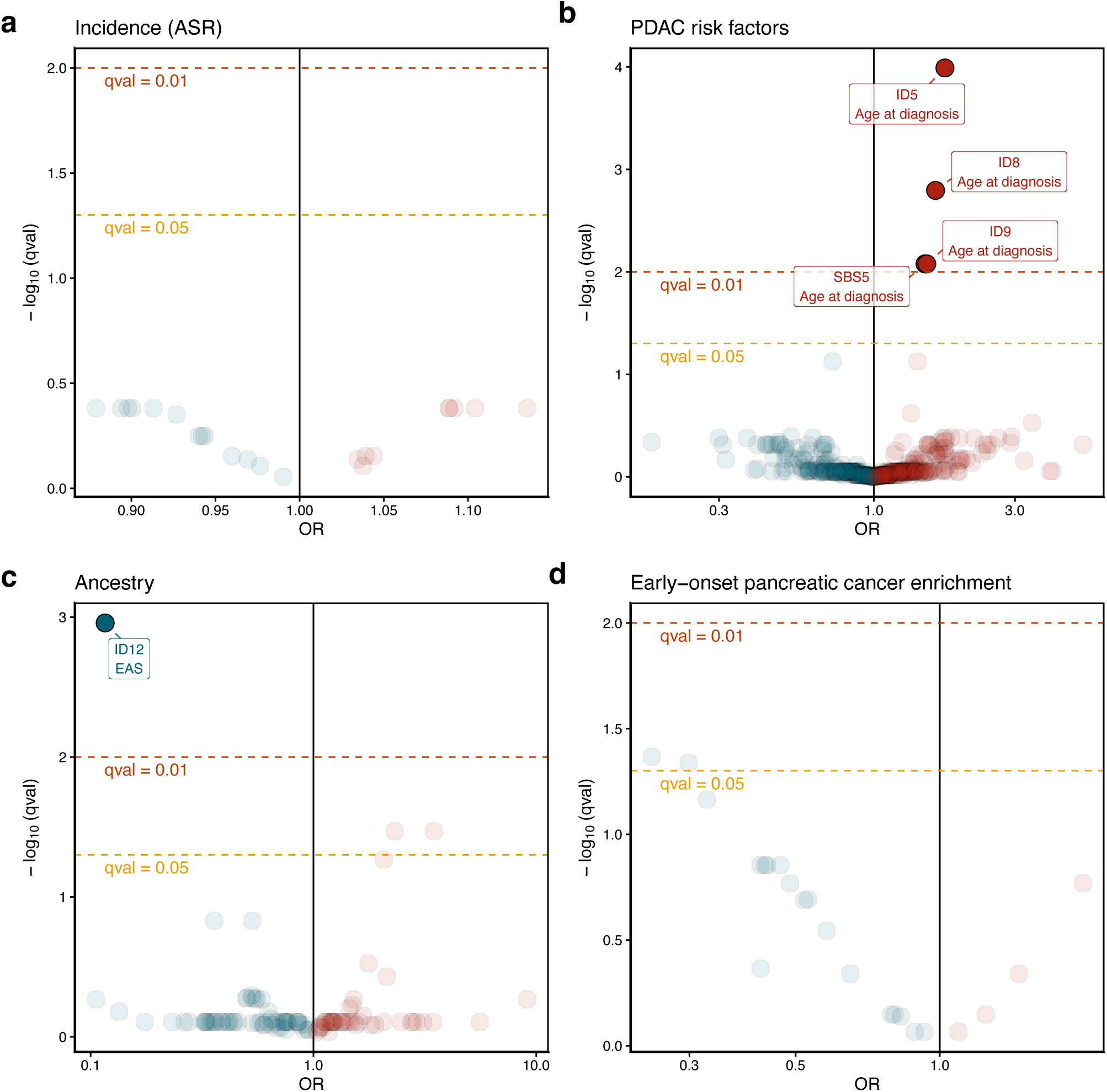
Association between mutational signatures and epidemiological data. **a)** Enrichment of mutational signature activities with pancreatic cancer age-standardized incidence rates (ASR). **b)** Associations of mutational signature activities with known or suspected pancreatic cancer risk factors and clinical factors. **c)** Enrichment of mutational signature activities across genetic ancestries. **d)** Enrichment of mutational signature activities in early-onset pancreatic cancers (age at diagnosis < 50 y.o.) compared to late-onset. Multivariate logistic regression models adjusted for age at diagnosis (all panels except d), sex (all panels), tumour purity (all panels) and country (only for panels b and d) were used to test for association. Firth’s bias-reduced logistic regression was applied when complete or quasi-complete separation occurred or when standard models did not converge. P values were corrected using the Benjamini– Hochberg procedure and reported as q values; tests with significant differences (q < 0.01) are labelled.

## Discussion

The mutational signature analysis of 495 pancreatic cancer genomes from nine countries has revealed a landscape which is remarkably consistent across all the countries studied. This landscape is different from that previously reported for pancreatic cancer genomes due to the presence of an additional mutational signature, SBS111, which is present in almost all cases and contributes just over a quarter of the total mutation burden^2,17,18^. It is known that including cases with DNA repair deficiency in the extraction process impairs the ability to extract SBS111, which potentially explains why it has not been reported previously^4,22^. The aetiology of SBS111 is unknown, but it is speculated to be caused by infidelity in DNA repair due to the association with indel burden, the overlap in SBS96 contexts with multiple DNA repair deficiency signatures and the similarity in the topographical features of C>A mutations compared to those caused by POLD1 deficiency^22,25^. This appears to be distinct from complete DNA repair deficiency, as these cancers do not show hypermutation or any canonical defects in genes known to cause DNA repair deficiencies. Therefore, it can be hypothesised that SBS111 represents a less severe impairment of multiple DNA repair pathways compared to the more severe canonical defects in a specific pathway. Regardless, the almost ubiquitous presence and high relative burden of SBS111 make its role in pancreatic cancer development an interesting avenue for future studies.

We find that no mutational signatures can be associated with differences in pancreatic cancer incidence rates. Neither do we identify any mutational signatures which can be conclusively linked to an exogenous exposure, which is consistent with previous whole-genome sequencing studies^17,18,26^. The simplest explanation for this is that the pancreas simply does not have the opportunity to acquire mutations from exogenous exposures directly and is only susceptible to agents which promote cancer via non-mutagenic mechanisms (promotional carcinogens). We know that such exposures exist from mouse studies, where exposure to human carcinogens did not produce a mutational signature for the majority of compounds tested^27^. It is also suspected that promotional carcinogens contribute to the much more extreme variation seen in esophageal squamous cell carcinoma incidence rates, where again no difference in the mutational signature landscape could be detected^7^. In the pancreas it has been hypothesised that tobacco smoking stimulates inflammation and fibrosis which promotes the development of pancreatic cancer^28^. If environmental exposures other than tobacco are present and act as promotional carcinogens in a similar way, then these would be difficult to detect using the mutational epidemiology approach unless they result in variations in driver mutation frequency. Such variations can arise when the clonal dynamics of tissues are influenced by environmental or lifestyle factors^29,30^, but we did not observe any such variation in this study.

In summary we have shown that the mutational signature landscape of pancreatic cancers is consistent across countries and does not reveal the presence of known or novel mutagenic exposures. Indeed, pancreatic cancers are unique among all the cancer types studied using the Mutographs study design yielding neither associations between mutational signatures and risk factors nor any signatures which display geographic variation^3,4,6,7^. These results also imply that the underlying causes of pancreatic cancer that remain to be discovered are unlikely to be strongly mutagenic and may instead be acting as promotional carcinogens that promote clonal development. The dataset we have generated will also be useful for future studies, being the most geographically and ethnically diverse source of whole-genome sequencing data for pancreatic cancers currently available.

## Methods

### Recruitment of cases and informed consent

Recruitment was coordinated by The International Agency for Research on Cancer (IARC/WHO) including 15 collecting centres across nine countries (**Supplementary Tables 1 & 22**). Eligible patients were ≥18 years of age (ranging from 28 to 89 years with a mean of 65 years and standard deviation of 10 years) and a confirmed diagnosis of primary pancreatic cancer with no prior treatment. Informed consent was obtained for all participants. Patients were excluded if they had any condition that could interfere with their ability to provide informed consent or if there were no means of obtaining adequate tissues or associated data as per the protocol requirements. Ethical approvals were first obtained from each Local Research Ethics Committee and Federal Ethics Committee when applicable, as well as from the IARC Ethics Committee.

### Biospecimens and data collection

Dedicated standard operating procedures, following guidelines from the International Cancer Genome Consortium (ICGC), were designed by IARC/WHO to select appropriate case series with complete biological samples and exposure information as described previously^5,7^. In brief, for all case series included anthropometric measures were taken, together with relevant information regarding medical and familial history. All biological samples from retrospective cohorts were collected using rigorous, standardized protocols and fulfilled the required standards of sample collection defined by the IARC/WHO for sequencing and analysis. Potential limitations of using retrospective clinical data collected using different protocols from different populations were addressed by a central data harmonization to ensure a comparable group of exposure variables (**Supplementary Table 22**). All patient-related data were pseudonymized locally through the use of a dedicated alpha-numerical identifier system before being transferred to the IARC/WHO central database.

### Expert pathology review

Original diagnostic pathology departments provided diagnostic histological details of contributing cases through standard abstract forms, together with a representative hematoxylin–eosin-stained slide of formalin-fixed paraffin-embedded tumor tissues whenever possible. The IARC/WHO centralized the entire pathology workflow and coordinated a centralized digital pathology examination of frozen tumor tissues collected for the study, as well as formalin-fixed paraffin-embedded sections when available, via a web-based report approach and a dedicated expert panel, following standardized procedures as described previously^7,31^. A minimum of 50% viable tumor cells was required for eligibility for whole-genome sequencing.

### Laser capture microdissection and DNA extraction

A total of 979 patients with primary pancreatic cancer were enrolled in the study, including biological samples for 862 cases and sequencing data (FASTQ files) for 30 cases from Japan and 87 cases from Canada. Of these, 627 proceeded while 352 samples (41%) were excluded prior to sequencing due to insufficient viable tumour cells (pathology level) and/or inadequate DNA (tumour or germline). In order to maximise tumour cell content LCM was performed on 369 samples as previously described^31^. Extraction of DNA from fresh frozen tumors and matched blood/normal samples was centrally conducted at IARC/WHO with the exception of 110 cases from Canada and all 104 cases from USA. Germline DNA was extracted from either blood samples (580 cases) or normal tissues (47 cases) using previously described protocols and methods^5,7^.

### Whole-genome sequencing

In total 510 cases were received at the Wellcome Sanger Institute for whole genome sequencing. Fluidigm single nucleotide polymorphism (SNP) genotyping with a custom panel was performed to ensure that each pair of tumour and matched normal samples originated from the same individual. Whole-genome sequencing (150 bp paired end) was performed on the Illumina NovaSeq 6000 platform with target coverage of 40X for tumours and 20X for matched normal tissues. From this point both the 510 internally sequenced samples and the 117 externally sequenced samples were analysed using the same pipeline. Reads were aligned to the GRCh38 human reference genome using Burrows-Wheeler-MEM (v0.7.16a and v0.7.17)^32^. Post-sequencing quality control metrics were applied for total coverage, evenness of coverage and contamination. Cases were excluded if coverage was below 30X for tumour or 15X for normal tissue. For evenness of coverage, the median over mean coverage score was calculated. Tumours with median over mean coverage scores outside the range of values determined by previous studies to be appropriate for whole-genome sequencing (0.92–1.09) were excluded^33^. Conpair^34^ was used to detect contamination. Cases were excluded if the estimated contamination was greater than 3%. Samples with <1,000 total somatic SBS variants were also excluded. A total of 533 cases of pancreatic cancer and matched normal tissue passed all criteria.

### Somatic variant calling

Variant calling was performed using the standard Sanger bioinformatics analysis pipeline (https://github.com/cancerit). Copy number profiles were determined first using the algorithms ASCAT^35^ and BATTENBERG^36^, where tumour purity allowed. Single nucleotide variants (SNVs) were called with cgpCaVEMan^37^, indels were called with cgpPINDEL^38^, and structural rearrangements were called using BRASS (https://github.com/cancerit/BRASS). CaVEMan and BRASS were run using the copy number profile and purity values determined from ASCAT where possible (complete pipeline, n = 471). Where tumour purity was insufficient to determine an accurate copy number profile (partial pipeline, n = 61), CaVEMan and BRASS were run using copy number defaults and an estimate of purity obtained from ASCAT/BATTENBERG. Finally, for a single case which had no large CNs (copy number normal pipeline), CaVEMan and BRASS were run using copy number defaults and an estimate of purity calculated by the median variant allele frequency (VAF) of IDs multiplied by two. For SNVs, additional filters on ASRD (read length-adjusted alignment score of reads showing the variant allele) and CLPM (median number of soft-clipped bases in variant supporting reads) (ASRD ≥ 140 and CLPM = 0) were applied to remove potential false positive calls. A second variant caller, Strelka2^39^, was run for SNVs and indels as consensus variant calling was previously shown to eliminate algorithm-specific artefacts and to generate highly reproducible mutational spectra compared to using a single variant calling algorithm^7^. Only variants called by both the Sanger variant calling pipeline and Strelka2 were included in subsequent analysis.

### Driver mutations

Driver mutations were called as previously described^3^. In brief, dNdS (https://github.com/im3sanger/dndscv)^40^ was used to identify genes under selection. A gene was considered under selection if significant in either the whole-genome (q value < 0.01) or in a restricted panel of 369 genes (q value < 0.05) previously confirmed to be under selection in cancer^40^. All variants in genes identified as being under selection in the global dNdS result and/or in the 369 cancer gene panel were then classified as likely drivers if they met any of the following criteria: (1) truncating mutations in genes annotated as tumor suppressors, (2) mutations annotated as likely or known oncogenic in MutationMapper, (3) truncating variants in genes with selection (q < 0.05) for truncating mutations assumed to be tumor suppressors and thus likely drivers, (4) missense variants in all genes under positive selection and with dN/dS ratios for missense mutations above five (assuming four of every five missense mutations are drivers) classified as likely drivers or (5) in-frame indels in genes under significant positive selection for in-frame indels. Mode of action was annotated using the Cancer Gene Census (https://cancer.sanger.ac.uk/census) and Cancer Genome Interpreter (https://www.cancergenomeinterpreter.org)^41^. Missense mutations were assessed using MutationMapper (http://www.cbioportal.org/mutation_mapper)^42^.

### Extraction of mutational signatures

Mutational matrices were generated using SigProfilerMatrixGenerator (https://github.com/AlexandrovLab/SigProfilerMatrixGenerator)^43^ with default options (v1.2.26). Mutational signatures were primarily extracted using SigProfilerExtractor (https://github.com/AlexandrovLab/SigProfilerExtractor)^20^ based on non-negative matrix factorization and then validated using mSigHdp (https://github.com/steverozen/mSigHdp)^19^, based on hierarchical Dirichlet process mixture models. SigProfilerExtractor (v1.1.24) was run using nndsvd_min initialization (NMF_init=‘nndsvd_min’) for 1–20 signature solutions and 500 non-negative matrix factorization (NMF) replicates. For SBS, mutational signatures were extracted in SBS96, SBS288 and SBS1536 contexts. All results were similar (**Supplementary Note**). Indel signatures were extracted using SigProfilerExtractor for ID83 contexts. Signatures were extracted using mSigHdp with the recommended parameters and no hierarchy for SBS96 and ID83 contexts only as mSigHdp has not been benchmarked for extended contexts. Signature results were similar from both algorithms with a full comparison available in the Supplementary Note.

### Decomposition of mutational signatures

The extracted *de novo* mutational signatures were decomposed into COSMIC v3.6 reference signatures using SigProfilerAssignment (v0.1.4, https://github.com/SigProfilerSuite/SigProfilerAssignment)^44,45^. To optimise the decomposition, the exclude_signature_subgroups parameter was used to exclude artefact signatures, ultraviolet signatures, lymphoid signatures, mismatch repair deficiency signatures, polymerase deficiency signatures, homologous recombination deficiency signatures, base excision repair deficiency signatures, and treatment signatures. For SBS signatures decompositions were performed for SBS96, SBS288 and SBS1536 extractions and only reference signatures which were present in all three optimised decompositions were accepted into the final decomposition result (**Supplementary Note**).

### Attribution of mutational signatures

The activities of *de novo* and COSMIC mutational signatures were attributed using MSA (v2.0, https://gitlab.com/s.senkin/MSA) as described previously with the optional parameter params.no_CI_for_penalties = false enabled^23^. For COSMIC attributions, the panel consisted only of COSMIC reference signatures which were identified in the decomposition stage. MSA calculates confidence intervals for each attribution and produces a pruned attribution result where any signature activity with a lower confidence limit equal to 0 is removed. Unpruned results were used for the final analysis based on simulation results which showed that pruning the results is too conservative in this cancer type (Supplementary Note). ID83A and ID83B (primarily composed of ID1 and ID2 respectively) were deprioritised for optimal penalty selection during attribution of the *de novo* indel signatures. This is due to ID1 (and ID2 to a lesser extent) being a component in all the extracted *de novo* indel signatures leading to difficulties in attribution and optimal penalty selection. It is not possible to achieve a penalty with 95% specificity for ID83A and ID83B without significant loss of sensitivity to all signatures. Since ID1 and ID2 represent the slippage of the nascent (ID1) or template (ID2) strands during DNA replication and are known to be active in almost all cancer samples this is unlikely to have resulted in false positives^2^. As the COSMIC reference signatures do not contain these residual ID1 and ID2 components, this was not an issue for the COSMIC attributions.

### Simulated data sets

A simulated dataset of 500 samples was generated to validate the attribution results. Simulated sample mutation burdens were drawn from a uniform distribution over the range 1,000–5,000 mutations. Signature exposure proportions were sampled from a Dirichlet distribution with concentration parameters proportional to those observed in the real dataset (mean relative signature attribution × 10), resulting in variability comparable to that of the real data (**Supplementary Note**). These exposure proportions were scaled by the corresponding mutation burdens to obtain the expected mutation counts (λ) for each SBS96 context. To account for stochastic variation, final mutation counts were then drawn from a Poisson distribution with the corresponding λ parameters.

### Evolutionary analysis

Of the 495 cases used in the final analysis, 435 had Battenberg results available. In order to identify the clonal structure of each sample, the data was first pre-processed using dpclust3p (v1.0.8, https://github.com/Wedge-lab/dpclust3p) and used as input for DPClust (v2.2.8, https://github.com/Wedge-lab/dpclust)^36^. MutationTimeR (v1.00.1, https://github.com/gerstung-lab/MutationTimeR)^46^ was used to assign somatic mutations to a clonality category (early clonal, late clonal, subclonal or unspecified). MSA (v2.0)^23^ was then used to assign signature activities to each category. Unpruned attributions were used for final results. For each signature, differences between paired early- and late-clonal activities were assessed using the Wilcoxon signed-rank test, and p-values were adjusted for multiple testing using the Benjamini–Hochberg procedure. Adjusted p-values are reported as q-values.

### Ancestry analysis

Genetic ancestry was inferred from HapMap single-nucleotide polymorphisms (SNPs; n = 956,073) extracted from whole-genome sequencing–derived genotypes for 533 pancreatic cancer cases. Variant-level quality control was performed using PLINK v1.9b, excluding SNPs with minor allele frequency (MAF) < 1% in the cohort (n = 14,225), genotype missingness > 5% or deviation from Hardy–Weinberg equilibrium (p < 1 × 10^−8^; n = 1,621). After linkage disequilibrium pruning, 134,494 variants remained.

The filtered genotypes were merged with overlapping variants from the 1000 Genomes Project phase 3 reference panel (3,202 individuals representing five superpopulations). Principal component analysis (PCA) was performed on the combined dataset to characterize ancestry variation in the Mutographs pancreatic cancer cohort relative to reference populations. Global ancestry proportions were then estimated using ADMIXTURE with K = 5, yielding membership probabilities for African (AFR), Admixed American (AMR), East Asian (EAS), European (EUR) and South Asian (SAS) ancestries. Each sample was assigned to a single ancestry if its maximum membership probability exceeded an optimized threshold (θ = 0.7); samples not exceeding θ for any ancestry were classified as admixed (ADM). This categorical ancestry variable was used in downstream analyses analogously to the country-based stratification (**Supplementary Note**).

### Regressions and association of risk factors with signatures

For mutational signature–based analyses, signature attributions were dichotomized using unpruned attributions. Signatures were classified as absent when the attributed mutation count was 0 and present when the attribution was greater than 0. For signatures present in at least 50% of samples (all signatures except SBS17a, SBS17b, SBS18, SBS93, SBS103, ID17, and ID21), samples were instead dichotomized into values above or below the median number of attributed mutations. These binary variables were used as response variables in all regression analyses. Logistic regression was used throughout. When predictors exhibited complete or quasi-complete separation, or when standard logistic regression failed to converge, Firth’s bias-reduced logistic regression was applied using the logistf R package.

Associations between mutational signatures and pancreatic cancer incidence were assessed using age-standardized incidence rates (ASRs) obtained from the Global Cancer Observatory (GLOBOCAN, 2022) as explanatory variables. These models were adjusted for sex, age at diagnosis (continuous, scaled and centered) and tumour purity, and were performed at the sample level. Country- and ancestry-enrichment analyses were performed with each country or ancestry tested individually against all others as a binary explanatory variable. These models were similarly adjusted for sex, age at diagnosis and tumour purity. All regressions assessing associations with risk factors were adjusted for potential confounders by including sex, age at diagnosis, tumour purity and country as covariates. Early-onset pancreatic cancer was defined as age at diagnosis strictly below 50. Enrichment of individual signatures in early-onset disease was assessed using early-onset status as the explanatory variable and adjustment for sex, tumour purity and country. For signatures exhibiting limited zero inflation (absence in <10% of samples – that is SBS1, SBS40a, SBS111, ID1, ID2 and ID8), additional continuous regressions were performed for risk factor and country analyses, using quasi-Poisson generalized linear models to account for overdispersion in the signature attributions. These models were similarly adjusted for sex, age at diagnosis, tumour purity, and country for the risk factor analysis. For all incidence, country, ancestry, risk factor and early onset analyses, p values were adjusted for multiple testing using the Benjamini–Hochberg false discovery rate procedure. Adjustments were performed separately within each analytical context (for example, number of signatures multiplied by the number of risk factors tested for the risk factor analysis). Adjusted p values are reported as q values, with q < 0.01 considered statistically significant.

### Other statistical analysis

Statistical power for regressions involving risk factors and country of origin was estimated using simulation-based analyses. Fixed parameters were derived from the observed cohort, including the total number of cases, the mean and standard deviation of age at diagnosis, and the male-to-female ratio. Key parameters were varied across simulations, namely the proportion of individuals exposed to the risk factor or originating from a given country (5–50%) and the prevalence of the binary outcome (10– 50%). These simulations generated theoretical datasets reflecting plausible regression scenarios consistent with the empirical cohort. Binary outcomes were simulated using a logistic model. Odds ratios (ORs) for age and sex were randomly sampled from a uniform distribution between 0.5 and 1.5 using the base R function runif, while ORs for the risk factor or country of origin were systematically varied between 1.1 and 3.5. The linear predictor was defined as

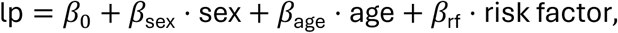

where *β*_0_ was set using the logit-transformed outcome prevalence (qlogis), and *β*_sex_, *β*_age_, and *β*_rf_ corresponded to the natural logarithms of their respective ORs. The binary response variable was then generated using a Bernoulli process (rbinom) with probabilities obtained from the logistic function (plogis) applied to the linear predictor.

For each combination of outcome prevalence, exposure proportion, and risk factor/country OR, 1,000 simulated datasets were generated. A logistic regression model including the risk factor (or country), age, and sex as covariates was fitted to each dataset. Statistical power was defined as the proportion of simulations in which the p-value for the risk factor or country variable was less than 0.05. Power estimation for incidence regressions followed an analogous simulation framework. Instead of a binary risk factor or country variable, an age-standardized rate (ASR) variable was assigned to individuals by random sampling from the empirical ASR values observed in the original dataset, with the constraint that each ASR value was represented in at least 10% of the cohort. Simulations were repeated 1,000 times for each combination of outcome prevalence and ASR OR. Power was calculated as the proportion of simulations in which the ASR term was statistically significant (p < 0.05).

## Data Availability

Whole-genome sequencing data, variant calling files and patient metadata have been deposited in the European Genome-Phenome Archive (EGA) associated with study EGAS00001002726. Aligned CRAM files for all cases are deposited in dataset EGAD00001016805, consensus SNV and indel variant calling files are in dataset EGAD00001016817 and patient metadata files are in dataset EGAD00001016816. The human reference genome used for alignment is available at ftp://ftp.sanger.ac.uk/pub/cancer/support-files/reference/GRCh38_full_analysis_set_plus_decoy_hla.fa. All other data are provided in the accompanying Supplementary Tables.

## Code availability

All algorithms used for data analysis are publicly available with versions and repositories noted in the methods and in the accompanying reporting summary. Code for figure generation, regression, ancestry analysis and power calculations is available at https://gitlab.com/Mutographs/mutographs-pancreas.

## Acknowledgements

The authors thank the IARC General Services, including the Laboratory Services and Biobank team led by Z. Kozlakidis and the Section of Support to Research overseen by C. Mehta; L. O’Neill, K. Roberts, K. Smith, S. Austin-Guest and the staff of Sequencing Operations at the Wellcome Sanger Institute for their contribution to sequencing; the work of all other collaborators in the Mutographs project who participated in the recruitment of patients in all centres; and all the patients involved in this study and their families. The authors also thank Maggie Blanks for her contributions as patient advocate during the Mutographs project. This work was delivered as part of the Mutographs team, supported by the Cancer Grand Challenges partnership funded by Cancer Research UK (C98/A24032). Work performed at the Sanger Institute was supported by the Wellcome Trust grants 206194 and 220540/Z/20/A. Work performed at UC San Diego was supported by Curebound Prize grant #Curebound_25CP01. Work performed by the Section of Support to Research at the IARC/WHO was supported by regular budget funding. Work at CNIO was supported by the Agencia Estatal de Investigación (AEI/10.13039/501100011033), Ministerio de Ciencia, Innovación y Universidades, and co-funded by the European Regional Development Fund (ERDF-EU; PID2024-158474NA-I00). M.D.-G. was awarded a fellowship within the “Generación D” initiative, Red.es, Ministerio para la Transformación Digital y de la Función Pública, for talent attraction (C005/24-ED CV1), funded by the European Union NextGenerationEU funds, through PRTR. This work was supported by grants from Practical Research for Innovative Cancer Control from the Japan Agency for Medical Research and Development (AMED) (26ck0106162h0001 to T.S.). The views expressed in this article by authors identified as IARC/WHO personnel are their own and do not necessarily represent the decisions, policies, or views of the IARC/WHO. The designations employed and the presentation of the material in this publication, in particular in Fig.1, do not imply the expression of any opinion whatsoever on the part of the authors or their institutions concerning the legal status of any country, territory, city, or area or of its authorities, or concerning the delimitation of its frontiers or boundaries. For the purpose of Open Access, the author has applied a CC BY public copyright licence to any Author Accepted Manuscript version arising from this submission. We remember Dr. Gloria Petersen, who contributed to the collaboration and inclusion of cases from the USA; this work honours her passion for science and her kind and generous spirit.

## Author Contributions

S.M., T.C., J.P., H.Z., C.N., S.F., P.G.-G., V.G., C.L., S.C., Se.S., G.T.T., A.P.L. and M.D.-G. performed data analysis. B.A.-A designed and oversaw the pathology workflow. B.A.-A., Ar.N. and St.S. performed pathology review. P.C., Ar.N., C.C. and I.L. performed sample manipulation. A.F.-I., R.M.R., D.P.G., P.A.-P., A.B.O., A.d.M., A.M.P.J., M.P.C., F.J.F.C., V.D., S.O., T.S., Sa.S., R.M., S.N., Ah.N., F.S., B.Ś., J.S., K.R.-P., J.L., I.H., P.Z., A.M., O.S., D.Z., S.O.A., A.L.O., S.G. and J.M.W. contributed to patient and sample recruitment. L.H., A.C.d.C. and S.P. managed the scientific project. L.B.A., P.B. and M.R.S. conceived, designed and supervised the study. S.M. and T.C. jointly contributed and were responsible for overall scientific co-ordination. S.M., T.C. and M.R.S. wrote the manuscript. All authors reviewed the manuscript and approved the final version.

---

**Extended Data Fig. 1.**
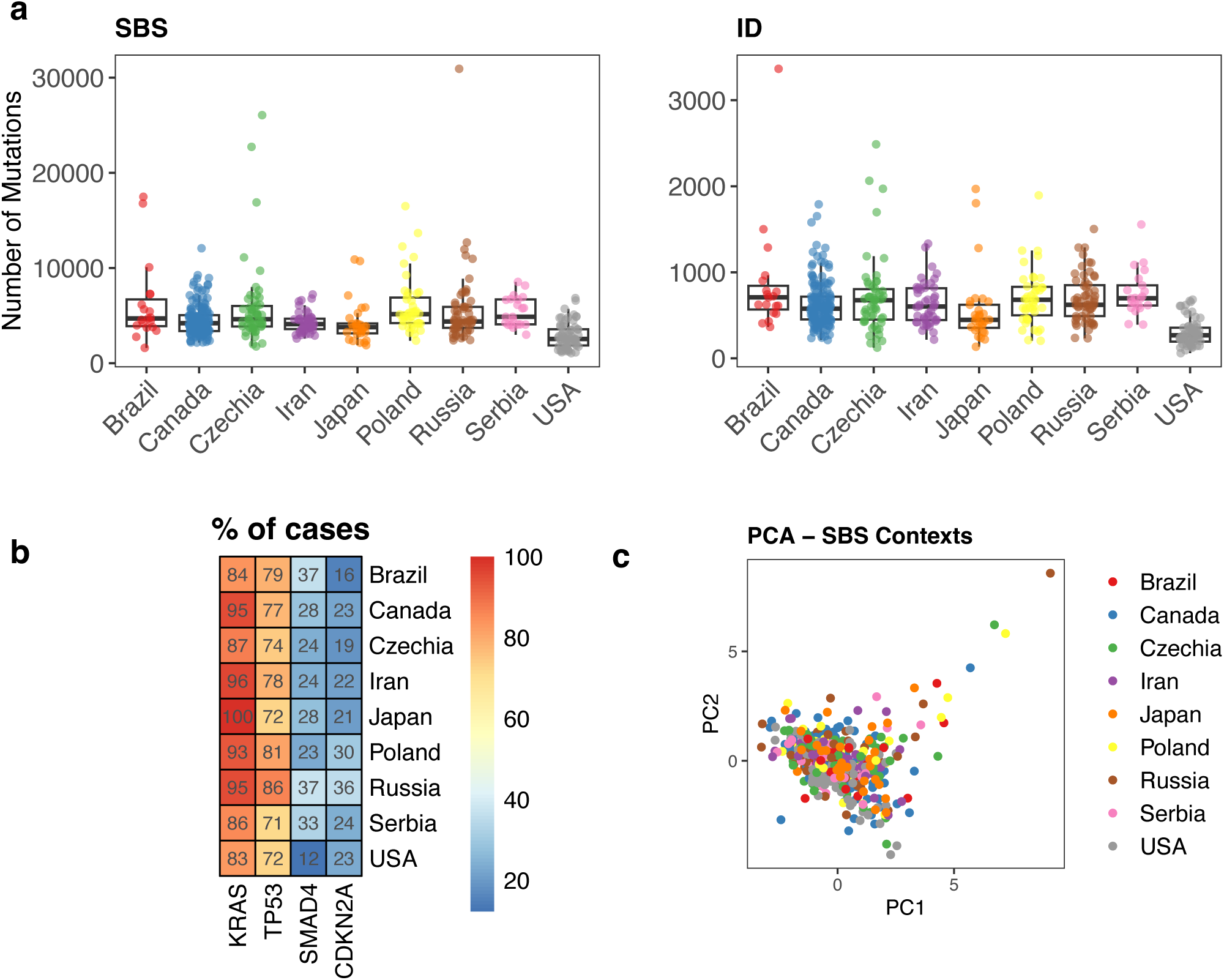
Mutation burdens and driver mutations in pancreatic cancers. **a)** Total SBS and ID mutation burden in 495 pancreatic cancers from nine countries. **b)** Frequency of driver mutations in pancreatic cancers from nine countries. Drivers present in >10% of pancreatic cancers were included. **c)** PCA analysis of relative SBS96 mutation counts on pancreatic cancers from nine countries. Box-and-whisker plots are in the style of Tukey. The line within the box is plotted at the median, while upper and lower ends indicate 25th and 75th percentiles. Whiskers show 1.5 × IQR.

**Extended Data Fig. 2.**
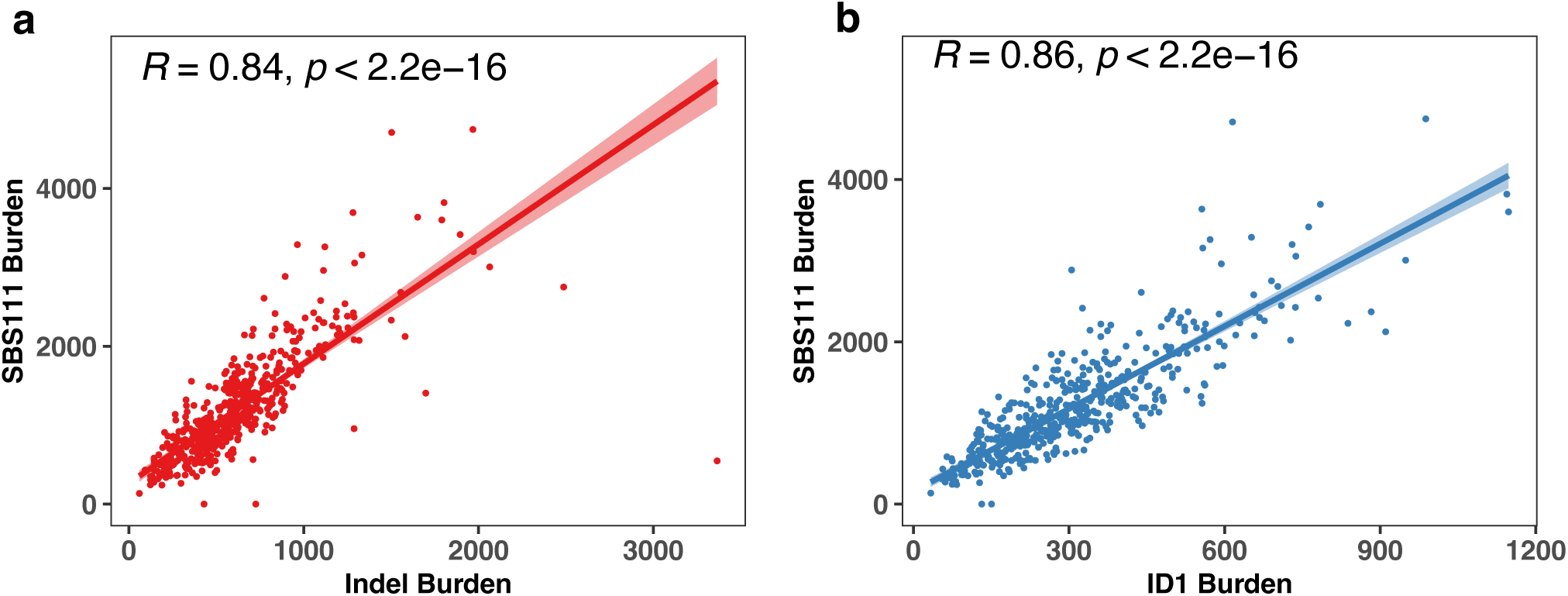
Correlation of Indel and ID1 burden with SBS111 burden. Correlation between **a)** total indel burden and **b)** ID1 burden with SBS111 burden. P values were calculated using Spearman’s rank correlation coefficient.

**Extended Data Fig. 3.**
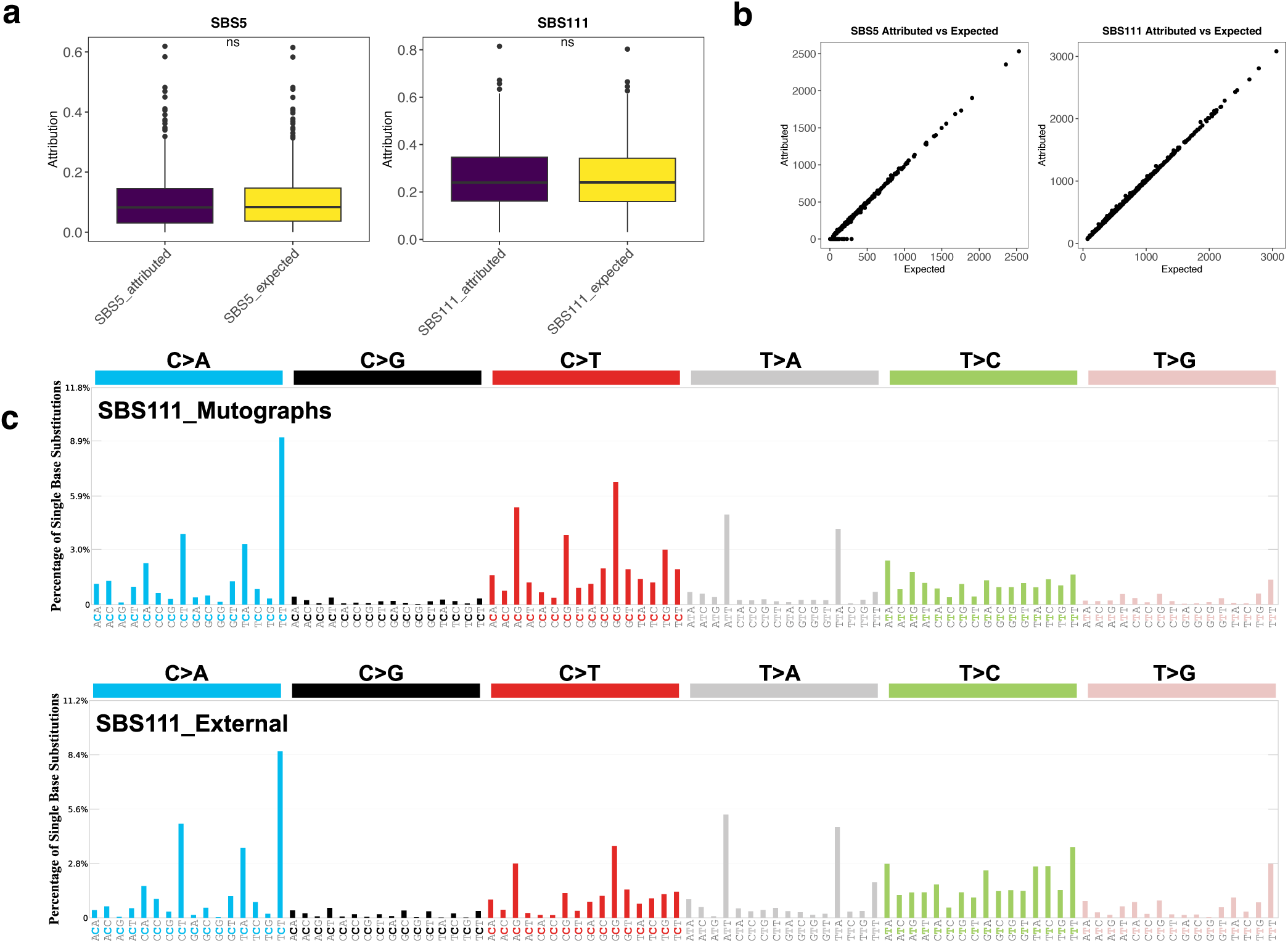
Validation of signature SBS111 in simulated and external datasets. **a)** comparison of expected vs attributed relative proportion of SBS5 and SBS111 in the MSA attribution. **b)** absolute expected vs attributed mutation burden of SBS5 and SBS111 in the MSA attribution. **c)** SBS111 signature extracted from the Mutographs (495 cases) and external (318 cases) datasets. Box-and-whisker plots are in the style of Tukey. The line within the box is plotted at the median, while upper and lower ends indicate 25th and 75th percentiles. Whiskers show 1.5 × IQR, and values outside it are shown as individual data points

