## Supplementary Note for "Mutational signature landscape of pancreatic cancer genomes from nine countries"

**Supplementary information inventory**

**This file contains the following:**

Supplementary Note

Supplementary References

Supplementary Figures

#### SUPPLEMENTARY NOTE

**Contents**

**1. Driver mutations**

1.1 dNdS results

1.2 Driver mutations

**2. Mutational signature analysis**

2.1 Samples excluded from mutational signature extractions

2.2 Extraction of *de novo* SBS mutational signatures with SigProfilerExtractor

2.3 Extraction of *de novo* signatures with mSigHdp

2.4 Decomposition to reference signatures

3. **Validation of mutational signature attribution**

3.1 Generation of simulated data

3.2 Attribution of simulated data

**4. Ancestry analysis**

4.1 Genetic ancestry distribution

**5. Regression analysis**

5.1. Supplementary ASR regressions

5.2. *De novo* regressions

5.3. Continuous regressions

5.4 Power calculations

**6. Germline analysis**

6.1 Germline variants in pancreatic cancers

6.2 Association between germline variants and mutational signatures

#### 1. Driver mutations

##### 1.1 dNdS results

A total of 38 genes were found to be under selection in pancreatic cancers, including 25 which were under selection in global dNdS and an additional 13 genes using restricted hypothesis testing (**Supplementary Note Table 1**). Overall, 24 of the 38 genes have been previously reported as cancer genes in pancreatic cancers and included the well-established canonical drivers *TP53*, *SMAD4*, *CDKN2A*, *KRAS* in addition to *TGFBR2*, *RNF43*, *ARID1A*, *ACVR1B*, *SF3B1*, *KDM6A*, *RBM10*, *ACVR2A*, *RB1*, *ATM*, *MAP2K4*, *SMAD3*, *GNAS*, *DUSP16* and *FBXW7* which were significant at the global level and *BCOR*, *BRAF*, *KMT2C*, *KMT2D* and *PBRM1* which were significant under restricted hypothesis testing [1-3]. An additional 10 genes; *ZFP36L2, PTEN, SOX9, PIK3R1*, *GPS2*, *MAP2K7*, *RREB1, KLF4, STK11* and *ELF3* have been reported as drivers in other cancer types or known to be under selection in human cancers[3,4]. The remaining 4 genes were *ERCC6L*, *PRSS1*, *MAP3K21* and *PDZD11*. The frequency of variants for each gene was less than 3% in the cohort overall, and these may represent novel rare driver genes in pancreatic cancer.

##### 1.2 Driver mutations

After classification (methods) a total of 1550 variants in 96 genes were annotated as likely driver mutations (**Supplementary Table 3**). The only genes mutated in more than 10% of samples overall were *KRAS* (455/495, 92%), TP53 (383/495, 77%), SMAD4 (131/495, 26%) and CDKN2A (119/495, 24%). There were no significant associations between driver frequency and country after false discovery rate correction (**Supplementary Fig. 1a, Supplementary Note Table 2**). For individual KRAS hotspot mutations there was an enrichment of G12V in cases from Japan, but this did not pass FDR correction ($p < 0.01$, $q = 0.17$, **Supplementary Fig. 1b, Supplementary Note Table 3**). There was no difference in the mutational signature landscape between different KRAS hotspot mutations (**Supplementary Fig. 1c**).

#### 2. Mutational signature analysis

##### 2.1 Samples excluded from mutational signature extractions

In total 533 cases passed all quality control metrics after whole genome sequencing. Of these, 38 cases were excluded from subsequent analysis including mutational signature analysis. The rationale behind excluding these is that previous Mutographs studies have demonstrated that including cases with high burden mutational signatures, like those which cause canonical DNA repair deficiencies, can prevent the extraction of certain mutational signatures such as SBS111 in microsatellite stable colorectal cancers[5]. Of the 38 excluded cases, 25 cases were excluded due to homologous recombination repair deficiency (HRD). 23 cases were classified as HRD using the Classifier of Homologous Recombination Deficiency (CHORD) tool (**Supplementary Note Table 4**)[6]. A further two cases were excluded due to their high p_hrd score (0.3 and 0.2 respectively) and flat SBS mutational spectra. 5 cases were excluded due to being classified as microsatellite unstable following review of mutational spectra (**Supplementary Fig. 2a**), of these most were also predicted to be MSI in the CHORD results (**Supplementary Note Table 4**) with a single case not called due to having less than 14,000 indels in repeat regions. 3 cases were excluded due to the presence of signatures resulting from chemotherapy treatments for previous cancers (**Supplementary Fig. 2b**). A single case each were found to have MUTYH (**Supplementary Fig. 2c**) or POLE mutations (**Supplementary Fig. 2d).** Two cases with high levels of SBS28 were also excluded (**Supplementary Fig. 2e)**. Finally, a single case with a mutational spectrum dominated by C[T>G]T mutations (**Supplementary Fig. 2f**) was excluded. This spectrum did not resemble any known COSMIC reference signature.

**2.2 Extraction of *de novo* mutational signatures with SigProfilerExtractor**

SBS signatures were extracted using SBS96, SBS288 and SBS1536 contexts. SBS288 and SBS1536 extend the SBS96 classification system in different ways, with SBS288 accounting for the SBS96 context in transcribed, untranscribed and intergenic regions of the genome and SBS1536 using a pentanucleotide context formed of the two flanking bases both 5' and 3' of the mutated base[7]. Extracting signatures using multiple contexts is beneficial, as prior to extraction it is not possible to assess which classification is the most effective at separating the mutational processes active in any given dataset. The results from SBS288 and SBS1536 were similar, with 9 SBS288 and 10 SBS1536 *de novo* signatures being extracted (**Supplementary Fig. 3**). Each of the SBS288 signatures had a similar counterpart in SBS1536 context with a cosine similarity above 0.97 (**Supplementary Note Table 5).** The additional SBS1536 signature represented an incomplete attempt to separate SBS17a from SBS17b, which does not affect the final panel of signatures attained as SBS17a would be decomposed from the combined signature regardless.

Extraction of doublet base substitution (DBS) signatures was attempted; however, the mutation burden was very low, ranging from 0 to 95 DBS variants with a median of just 9 variants per sample. Given that the standard DBS classification has 78 levels, the resulting matrices are too sparse to allow for reliable mutational signature extraction and attribution.

##### 2.3 Extraction of *de novo* signatures with mSigHdp

To ensure that we have captured the mutational signature landscape we ran a second mutational signature extraction using mSigHdp with the recommended parameters with no hierarchy[8]. mSigHdp extracted 9 SBS signatures (**Supplementary Fig. 4**) of which 7 were a close match to a corresponding SigProfilerExtractor signature with a cosine similarity above 0.95 (**Supplementary Note Table 5).** The remaining two signatures had lower cosine similarity to their SigProfilerExtractor counterparts, but this can be explained in both cases by incomplete separation of SBS1 in the SigProfilerExtractor *de novo* signature which is not present in the mSigHdp versions (**Supplementary Fig. 4, Supplementary Note Table 5).** mSigHdp extracted 7 Indel signatures which was the same number as extracted using SigProfilerExtractor (**Supplementary Fig. 5)**, of these 5 were a close match in both extractions with a cosine similarity above 0.97 (**Supplementary Note Table 6).** The remaining two signatures had a lower cosine similarity which was explained by the presence of peaks consistent with ID1 and ID2 in the SigProfilerExtractor versions but not the mSigHdp versions (**Supplementary Fig. 5, Supplementary Note Table 6).** Overall, the mutational signature extraction results were consistent between SigProfilerExtractor and mSigHdp.

##### 2.4 Decomposition to reference signatures

Decomposition was performed on the SBS96, SBS288, SBS1536 and ID83 SigProfilerExtractor *de novo* signatures (**Supplementary Note Table 7&8)**. The results for most SBS signatures were very similar but discrepancies were observed in 3 sets of equivalent signatures. A conservative approach was taken where only signatures which appeared in the final decomposition panel of all three extractions were taken forward to the final panel. This led to the exclusion of 6 signatures, SBS12, SBS34, SBS39, SBS40b, SBS102 and SBS112. SBS12 was present in 2 of the 3 decompositions and has previously been identified as an exposure in kidney and liver cancers from Japan[9]. The current study does include cases from Japan, but if SBS12 is allowed into the panel it is not attributed in any Japanese cases but rather randomly distributed amongst other countries. SBS39 was also present in 2 of 3 decompositions but was only attributed in a single case if allowed into the attribution panel. The remaining signatures were only present in the final signature panels from a single decomposition.

**3. Validation of mutational signature attribution**

**3.1 Generation of simulated data**

In order to provide a ground truth for validating the attribution of mutational signatures we generated a simulated dataset which is similar to the relative proportions of the signatures in the initial attribution (**Supplementary Fig. 6a**). When simulating the signature exposure proportions, using the mean relative signature attribution unadjusted resulted in a dataset where the majority of samples had very low or no SBS5, whereas multiplying this parameter by 10 gave sufficient variation to allow meaningful evaluation of the attribution process. The mutation burdens were sampled between 1000 and 5000 in line with the median burden of 4216 single base substitutions per pancreatic cancer genome.

**3.2 Attribution of simulated data**

The activities of each signature were attributed to the simulated dataset using MSA. This showed a difference between the average expected and attributed levels of SBS5 (11% expected, 6% attributed) and SBS111 (26% expected, 29% attributed) (**Supplementary Fig. 6b&c**). However, this was not the result of misattribution of SBS5 mutations to SBS111 but instead due to the process of pruning whereby MSA will remove individual signature attributions where the lower confidence interval is equal to 0. When assessing the unpruned results, the attributions performed better for both SBS5 (11% expected, 10% attributed) and SBS111 (26% expected, 26% attributed). All other signatures also showed good performance (**Supplementary Fig. 6d, Supplementary Note Table 9**). These results suggested that in some contexts, the pruning process of MSA can be too conservative and that using simulated datasets is a good way to detect this. On this basis the unpruned MSA attributions were used for all subsequent analysis.

#### 4. Ancestry analysis

##### 4.1. Genetic ancestry distribution

In total 423 samples were classified as EUR, 43 as EAS, 4 as AFR, 1 as SAS, 0 as AMR and 62 as ADM. Most countries were composed predominantly of individuals of European ancestry. Notable exceptions were Japan (100% EAS), Iran (98% ADM), and, to a lesser extent, Brazil (58% EUR, 37% ADM, and 5% EAS) (**Supplementary Fig. 7**).

#### 5. Regression analysis

##### 5.1 Supplementary ASR regressions

The national incidence rates for Iran and Brazil are the lowest of the countries included in this study. However, the cases collected from these countries were from urban industrialised cities which may have higher incidence rates compared to the national incidence. In order to exclude the possibility that this affected the results of the ASR regression analysis the analysis was performed without Iran and Brazil included. This showed a significant association of ID12 with ASR (**Supplementary Fig. 8, Supplementary Note Table 10**), however this is driven by the absence of ID12 in Japanese cases and does not hold when they are excluded. No other association was significant after FDR correction.

##### 5.2. *De novo* regressions

Regression analyses of the *de novo* mutational signatures were conducted using the same framework as that applied to the COSMIC signatures (see Methods). No *de novo* signature showed an association with ASR that could account for differences in incidence across countries (**Supplementary Note Table 11, Supplementary Fig. 9a**). Furthermore, no significant associations between *de novo* signatures and risk factors were observed after false discovery rate correction, with the exception of positive associations with age at diagnosis for SBS288C (a signature containing SBS5; OR = 1.87, q = 9.8 × 10−7) and ID83C (a signature containing ID1/5/8/9; OR = 1.94, q = 9.8 × 10−7) (**Supplementary Note Table 12, Supplementary Fig. 9b**).

Country-specific regression analyses identified several signatures depleted in the USA relative to all other countries, including SBS288D (OR = 0.19, q = 9.8 × 10−4), ID83A (OR = 0.14, q = 3.8 × 10−3) and ID83F (OR = 0.16, q = 1.2 × 10−3) (**Supplementary Note Table 13, Supplementary Fig. 9c**). By contrast, ID83D (an ID12-containing signature) was significantly enriched in the USA (OR = 5.9, q = 1.3 × 10−4), while showing strong depletion in Japan (OR = 0.013, q = 3.2 × 10−8). Consistent with the pattern observed for ID12, ID83D was also depleted in cases of East Asian ancestry (OR = 0.11, q = 5.6 × 10−4 – **Supplementary Note Table 14, Supplementary Fig. 9d**), largely reflecting its near absence in Japanese cases (presence in cases with East Asian ancestry in Japan: 1/29, 3.4%; with East Asian ancestry outside Japan: 11/13, 84.6%; with other ancestries: 418/453, 92.3%). Lastly, no *de novo* signatures exhibited a significant enrichment in early-onset pancreatic cases (**Supplementary Note Table 15, Supplementary Fig. 9e**).

**5.3. Continuous regressions**

As described in Methods, continuous regression models were additionally applied to assess associations between risk factors and signatures present in more than 90% of samples, namely SBS1, SBS40a, SBS111, ID1, ID2 and ID8. This approach did not materially increase statistical power, and only positive associations with age at diagnosis were identified for SBS1 (β = 0.092, q = 1.6 × 10−3) and SBS40a (β = 0.096, q = 4.3 × 10−4), consistent with the established clock-like behaviour of these signatures (**Supplementary Note Table 16, Supplementary Fig. 10a**). Country-specific regression analyses revealed depletion of several signatures in the USA relative to all other countries, including SBS1 (β = −0.33, q = 3.2 × 10−3), SBS111 (β = −0.47, q = 1.4 × 10−4), ID1 (β = −0.516, q = 3.8 × 10−5) and ID2 (β = −0.71, q = 2.8 × 10−3). By contrast, ID8 was significantly elevated in Brazil compared with the other countries included in the analysis (β = 0.476, q = 1.7 × 10−3) (**Supplementary Note Table 17, Supplementary Fig. 10b).**

##### 5.4 Power calculations

Power calculations were performed to calculate the size of effects detectable in this study. For ASR, simulations were performed using a range of signature prevalence (from 10 to 50% and higher) and randomly assigning ASR values drawn from the range of countries in the cohort. This showed that the minimal effect size required to reach 80% power is estimated to be between 1.3 and 1.7, indicating that relatively modest effects should be detectable (**Supplementary Fig. 11a**). For other risk factors a range of signature prevalences and risk factor proportions (from 5 to 50%, with the lowest comparable to the countries with the smallest number of cases and the highest reflective of common exposures such as tobacco and alcohol) were simulated. The minimal effect size required to reach 80% power was higher with odds ratios ranging from 1.7 to 2.6 (**Supplementary Fig 11b**). This becomes substantially higher when the risk factor is relatively rare (5%), or when both signature prevalence and risk factor proportions are low (10% each). Overall, the power calculations confirm that the analyses are sufficiently powered to detect moderate to strong effects with a limited ability to detect more modest effects.

#### 6. Germline analysis

##### 6.1 Germline variants in pancreatic cancers

The pathogenicity of germline variants was evaluated across a curated set of 189 cancer predisposition genes (CPGs). This gene set expanded upon 152 previously reported CPGs[10] by incorporating: 10 additional genes curated as CPGs by the ClinGen Hereditary Cancer Gene Curation Expert Panel; 10 genes with at least moderate evidence for tumour or neoplasia predisposition in the Gene Curation Coalition database (GenCC; accessed 1 August 2024, https://thegencc.org/); and 17 additional genes from the Cancer Gene Census germline catalogue (CGC-Germline; accessed 1 August 2024, http://cancer.sanger.ac.uk/census/). Genes were classified as PDAC-associated CPGs if at least moderate evidence supporting an association with pancreatic ductal adenocarcinoma (PDAC) was identified in ClinGen, GenCC or through literature review. 14 genes met these criteria: APC, ATM, BRCA1, BRCA2, CDK4, CDKN2A, MLH1, MSH2, MSH6, PALB2, PMS2, PRSS1, STK11 and TP53. Only variants present in one of these 14 genes were considered for analysis (**Supplementary Note Table 18**).

Germline single-nucleotide variants (SNVs) and small insertions or deletions (indels) were identified from whole-genome sequencing data generated from matched normal material. Variant calling was performed using Strelka2 with default parameters. Only PASS variants with DP ≥ 10 were considered for annotation. Variant interpretation was conducted according to the ACMG/AMP guidelines and updated ClinGen recommendations[11]. All variants detected in PDAC-associated CPGs were manually reviewed, applying ClinGen criteria specifications (CSpec) where available. For variants classified as variants of uncertain significance under the standard ACMG/AMP framework, a points-based classification approach was additionally applied[12]. Germline analysis of the 14 PDAC-associated CPGs was performed using WGS data from 533 patients with PDAC. Initial filtering identified 175 unique rare, non-benign variants according to ClinVar annotation, all of which underwent manual curation. Overall, 31 variants were classified as pathogenic or likely pathogenic (P/LP) across 8 PDAC-associated CPGs: *APC* (1 variant), *ATM* (9), *BRCA2* (11), *CDKN2A* (3), *MLH1* (1), *MSH6* (1), *PALB2* (2) and *PRSS1* (3).

In total, 54 cases (10.1%) carried at least one P/LP variant in a PDAC-associated CPG. This proportion decreased to 38 cases (7.7%) when restricting the analysis to cases retained for mutational signature analysis, after exclusion of DNA-repair-deficient tumours. Notably, 28 cases, including 24 in the signature-analysis subset, carried a pathogenic germline variant in PRSS1, a gene associated with hereditary pancreatitis. The recurrent PRSS1 c.47C>T (p.Ala16Val) variant was found in 26 of these cases, including 22 in the subset. The high prevalence of this variant within the cohort might be due to false-positive calls within PRSS1, a common issue likely caused by the strong sequence homology between the pseudo-gene PRSS3P3 and the protein-coding genes PRSS1 and PRSS2[13]. Accordingly, this variant was excluded from subsequent analyses. Following its removal, the number of cases carrying a P/LP variant in a PDAC-associated CPG decreased to 31 (5.8%) in the full cohort and 17 (3.4%) in the analysis subset, estimates that are more consistent with previously reported prevalences of germline pathogenic variants in PDAC[14]. In addition, 9 cases, all included in the signature-analysis subset, carried a different germline pathogenic variant in ATM (**Supplementary Note Table 19**).

##### 6.2 Association between germline variants and mutational signatures

To assess potential associations between germline pathogenic variants and mutational signatures, analyses were restricted to genes or gene groups harbouring pathogenic germline variants in at least 5 cases within the signature-analysis subset. These only included one PDAC-associated CPG, *ATM*, mutated in 9 cases. In addition, a complementary analysis was performed by aggregating all pathogenic variants across PDAC-associated CPGs, irrespective of the specific gene involved (17 cases).

Associations between germline pathogenic variant status and mutational signatures were evaluated using logistic regression models, following the same analytical framework as that used for risk-factor analyses. Dichotomised mutational signature exposure was modelled as the response variable, and the dichotomised presence of a pathogenic germline variant in each gene or gene group under investigation (*ATM*, any PDAC-associated CPG) was included as the explanatory variable. Models were adjusted for age, sex and tumour purity. Multiple testing correction was performed using the Benjamini–Hochberg false discovery rate procedure.

No statistically significant association between pathogenic germline variant carrier status in any analysed CPG and mutational signature exposure was observed after false discovery rate correction. This result is consistent with the composition of the analysed subset, from which DNA-repair-deficient cases had been excluded (**Supplementary Note Table 20**).


#### SUPPLEMENTARY FIGURES

**Supplementary Fig.1. Driver Mutations in pancreatic cancer genomes.** Enrichment of driver mutations in the four most frequently mutated driver genes (a) and KRAS hotspot mutations (b) across countries relative to all other countries. Analyses were based on dichotomized mutation status (presence/absence of driver mutations in KRAS, TP53, SMAD4, and CDKN2A; presence/absence of KRAS G12D, G12V, and G12R mutations). Multivariate logistic regression models adjusted for age at diagnosis, sex, and tumour purity were used to assess enrichment. Firth's bias-reduced logistic regression was applied when complete or quasi-complete separation occurred or when standard models did not converge. Points are coloured by odds ratio (OR), with point size indicating statistical significance. P values were corrected using the Benjamini–Hochberg procedure and reported as q values; tests with q < 0.05 are outlined in black. c) Average mutation burden of SBS mutational signatures in cases with specific KRAS hotspot mutations. All individual mutational signatures that contribute less than 5% on average are grouped in the ‘others’ category

**Supplementary Fig.2. Mutational spectra of cases excluded from mutational signature analysis.** Mutation spectra of cases excluded from further analysis due to a) microsatellite instability; b) chemotherapy treatment for previous cancers; c) MUTYH germline mutation; d) *POLE* mutation; e) high levels of SBS28 and f) hypermutator of unknown cause.

**Supplementary Fig.3: Single base substitution signatures extracted by SigProfilerExtractor.** All single base substitution (SBS) *de novo* signatures extracted in SBS96 (9 Signatures), SBS288 (9 signatures) and SBS1536 (10 signatures) format, shown side by side for comparison. An equivalent signature for SBS1536I was not extracted in SBS288 or SBS96 format.

**Supplementary Fig.4: Single base substitution signatures extracted by mSigHdp.** All single base substitution (SBS) *de novo* signatures extracted by mSigHdp.

**Supplementary Fig.5: Small insertion and deletion signatures extracted by mSigHdp.** All small insertion and deletion (ID) *de novo* signatures extracted by mSigHdp.

**Supplementary Fig.6. Validation of mutational signature attribution.** a) comparison of the simulated dataset versus the observed data from pancreatic cancers. b) expected vs attributed relative proportion of SBS5 and SBS111 in the pruned MSA attribution. c) absolute expected vs attributed mutation burden of SBS5 and SBS111 in the pruned MSA attribution. d) absolute expected vs attributed mutation burden of all COSMIC reference signatures in the unpruned MSA attribution. Box-and-whisker plots are in the style of Tukey. The line within the box is plotted at the median, while upper and lower ends indicate the 25th and 75th percentiles. Whiskers show 1.5 × IQR, and values outside it are shown as individual data points.

**Supplementary Fig.7. Genetic ancestry distribution.** Proportion of cases per country with African (AFR), East Asian (EAS), European (EUR), South Asian (SAS) or Admixed (ADM) ancestry.

**Supplementary Fig.8. Supplementary ASR Regressions.** a) Enrichment of mutational signature activities with pancreatic cancer age-standardized incidence rates (ASR) excluding cases from Iran and Brazil. b) proportion of ID12-positive cases across the studied countries in relation to pancreatic cancer incidence in each country. Dot size and color indicate the median attributed ID12 value per country. The regression line was fitted to country-level observations, assuming a linear association between the proportion of ID12-positive cases and ASR.

**Supplementary Fig.9. Regression analysis of de novo mutational signatures.** Enrichment of *de novo* mutational signature activities a) with pancreatic cancer age-standardized incidence rates (ASR); b) with known or suspected pancreatic cancer risk factors and clinical factors; c) across countries relative to all others; d) across genetic ancestries relative to all others; and e) in early-onset pancreatic cancers (age at diagnosis < 50 y.o.) compared to late-onset. Multivariate logistic regression models adjusted for age at diagnosis (all panels except e), sex (all panels), tumour purity (all panels) and country (only for panels b and e)) were used to test for association. Firth's bias-reduced logistic regression was applied when complete or quasi-complete separation occurred or when standard models did not converge. *P* values were corrected using the Benjamini–Hochberg procedure and reported as *q* values.

**Supplementary Fig.10. Continuous regression models.** Associations of COSMIC mutational signature activities a) with known or suspected pancreatic cancer risk factors and clinical factors; and b) across countries relative to all others. Multivariate Quasi-Poisson regression models adjusted for age at diagnosis, sex, tumour purity (all panels) and country (only for a) were used to test for association. Only non-zero-inflated signatures (> 90%) were assessed – specifically SBS1, SBS40a, SBS111, ID1, ID2 and ID8. P values were corrected using Benjamini-Hochberg procedure and reported as q values; tests with $q < 0.05$ are labelled.

**Supplementary Fig.11. Power calculations.** Power calculation results for (a) incidence-based regression analyses and (b) risk factor/country-based regression analyses as a function of effect size (odds ratio). In panel a, each curve corresponds to a different signature prevalence, ranging from 10% (darkest) to 50% (lightest). In panel b, each subpanel represents a distinct signature prevalence (10–50%), while curves within each subpanel correspond to different prevalences of the risk factor or country group, ranging from 5% to 50%. Each point represents the outcome of 1,000 simulations performed for a given combination of signature prevalence, risk factor/country prevalence, and effect size. Statistical power was estimated as the proportion of simulations yielding a significant association between the signature and either (a) incidence or (b) the corresponding risk factor/country. Accordingly, 80% power corresponds to at least 800 significant results across 1,000 simulations.

#### Supplementary Fig.1. Driver mutations in pancreatic cancer genomes

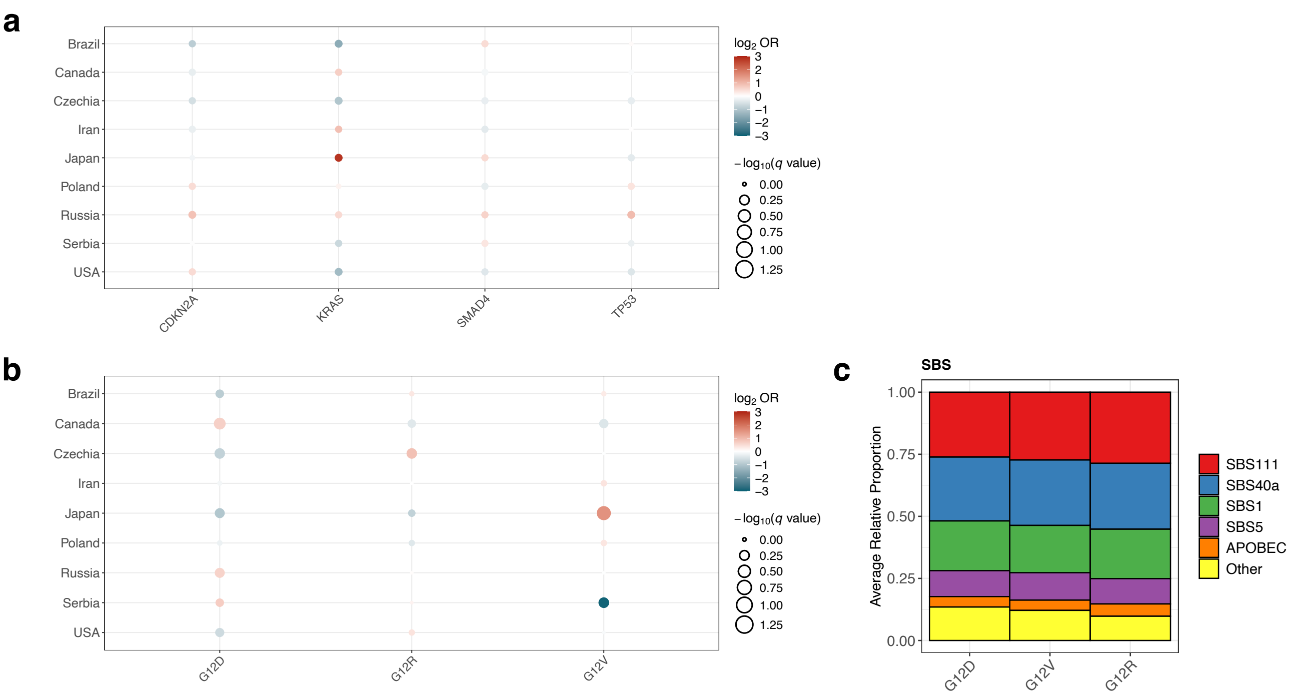

### Supplementary Fig.2. Mutational spectra of cases excluded from mutational signature analysis

a

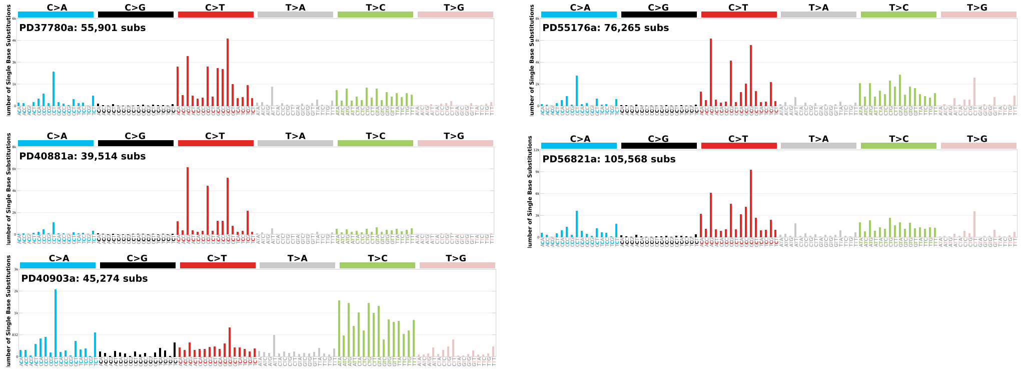


b

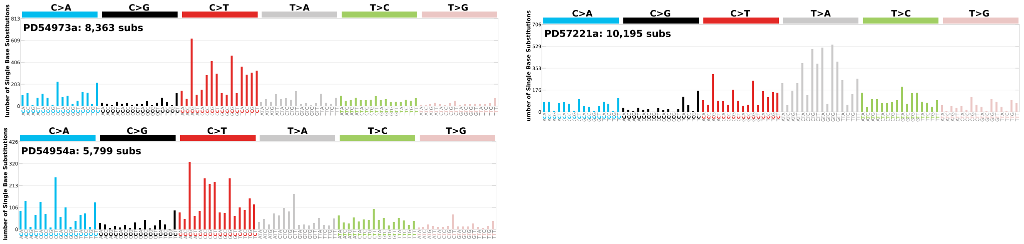


c

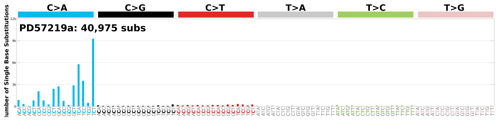


d

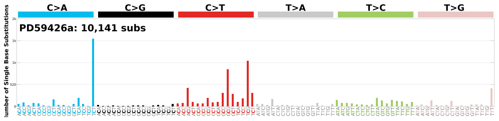


e

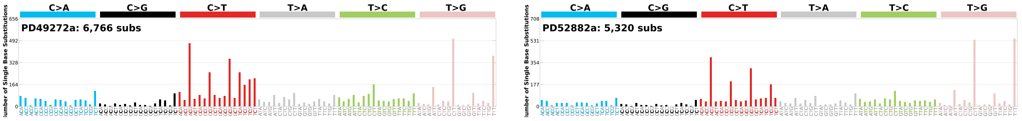


f

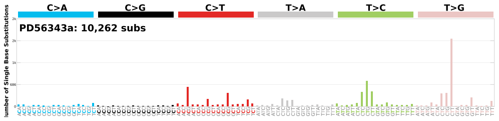

### Supplementary Fig.3. Single base substitution signatures extracted by SigProfilerExtractor

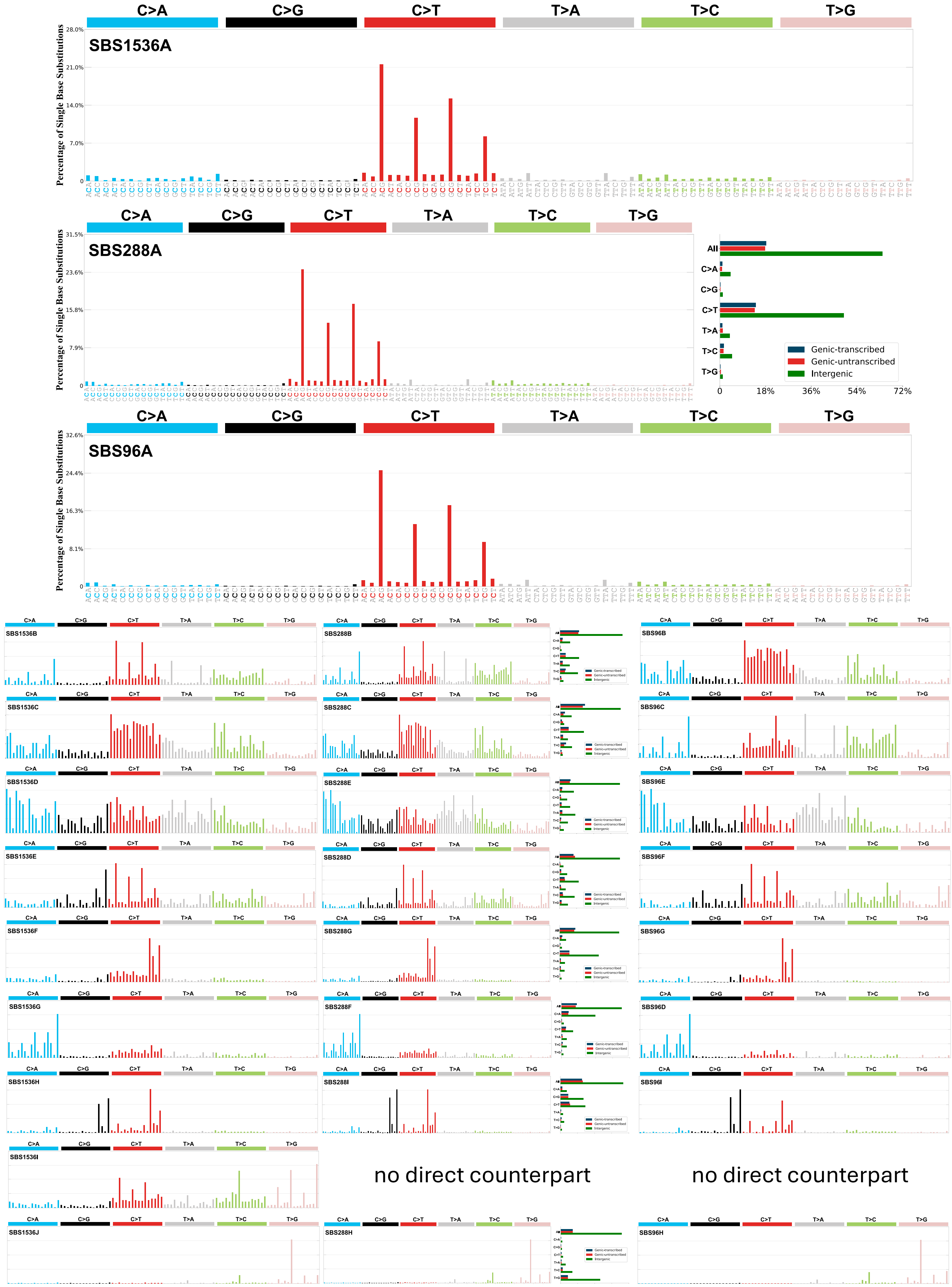

### Supplementary Fig.4. Single base substitution signatures extracted by mSigHdp

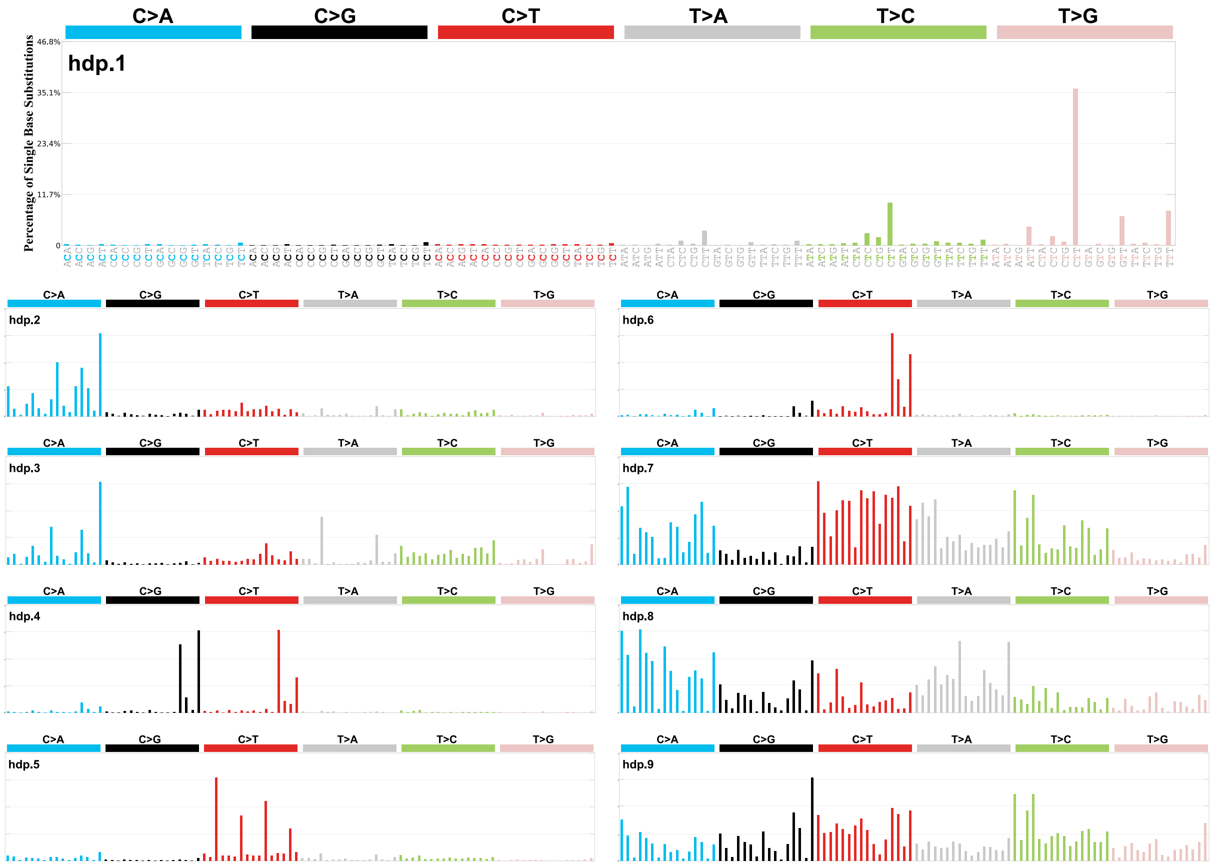

### Supplementary Fig.5. Small insertion and deletion signatures extracted by mSigHdp

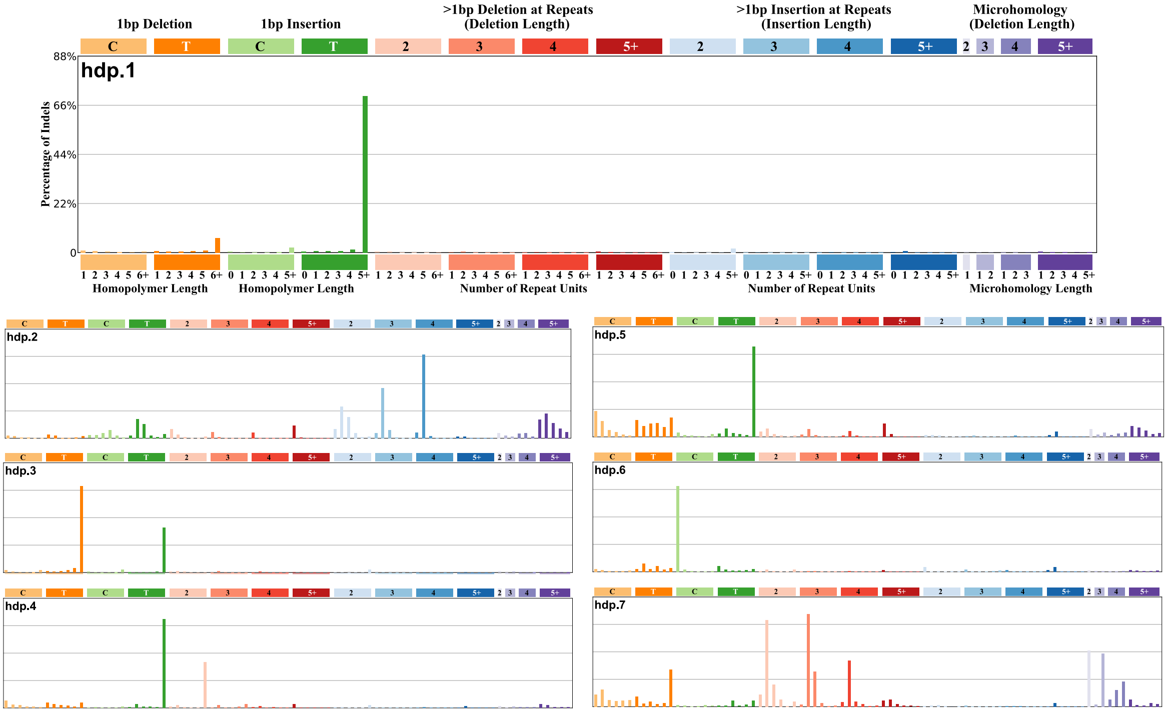

#### Supplementary Fig.6. Validation of mutational signature attribution

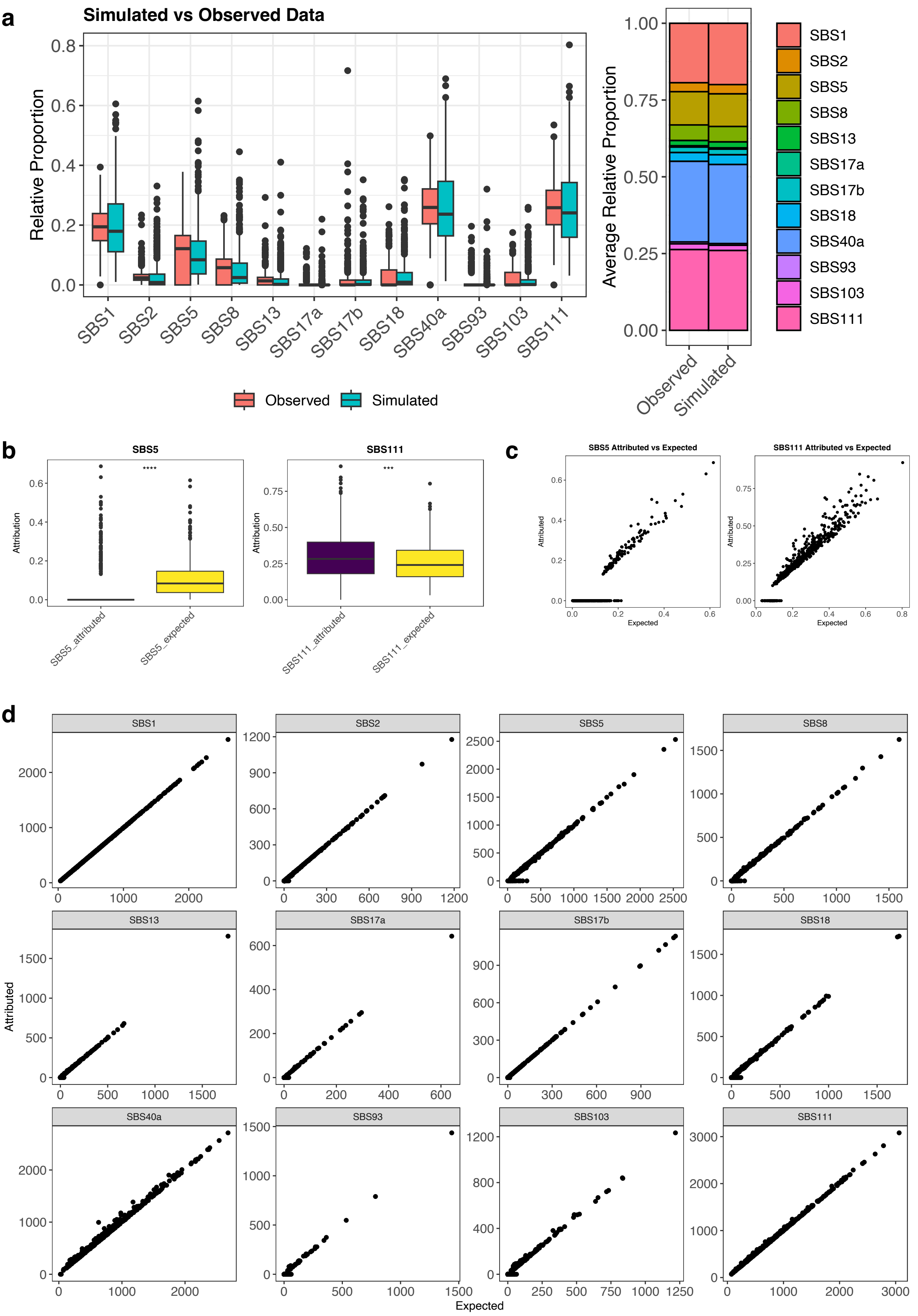

### Supplementary Fig.7. Genetic ancestry distribution

**a**

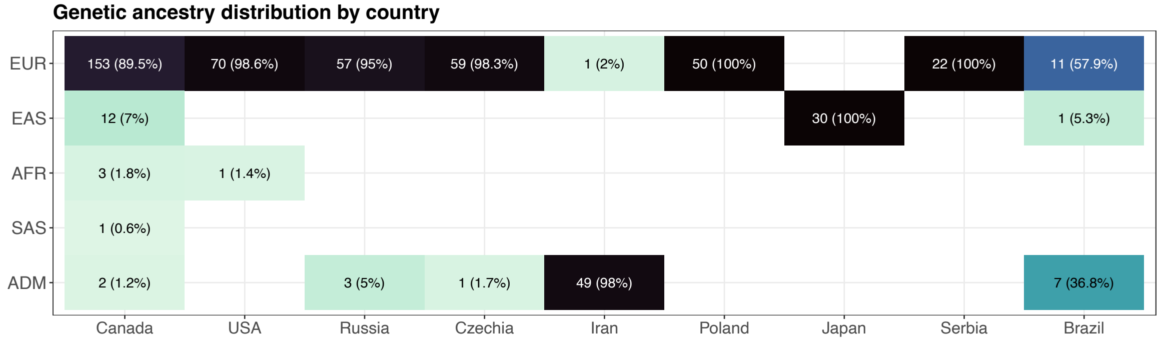

#### Supplementary Fig.8. Supplementary ASR regressions

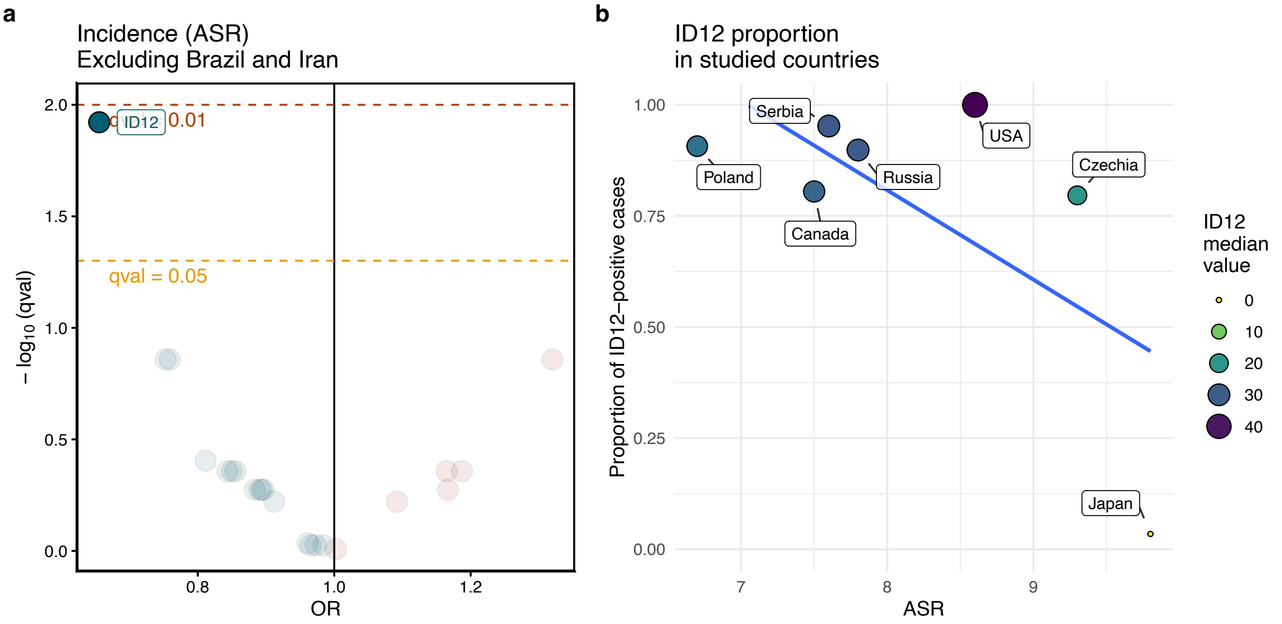

**Supplementary Fig.9. Regression analysis of de novo mutational signatures**

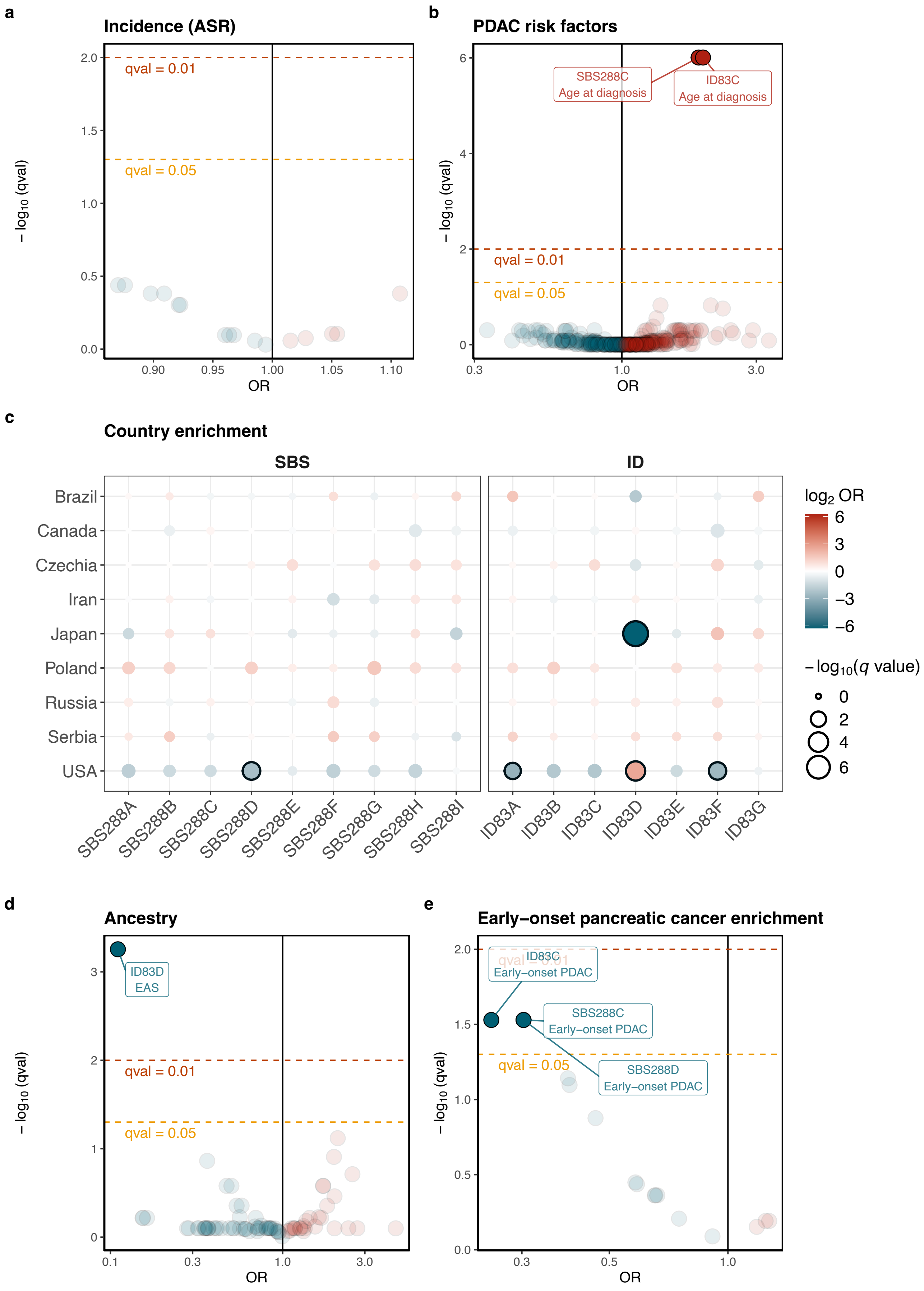

### Supplementary Fig.10. Continuous regression models

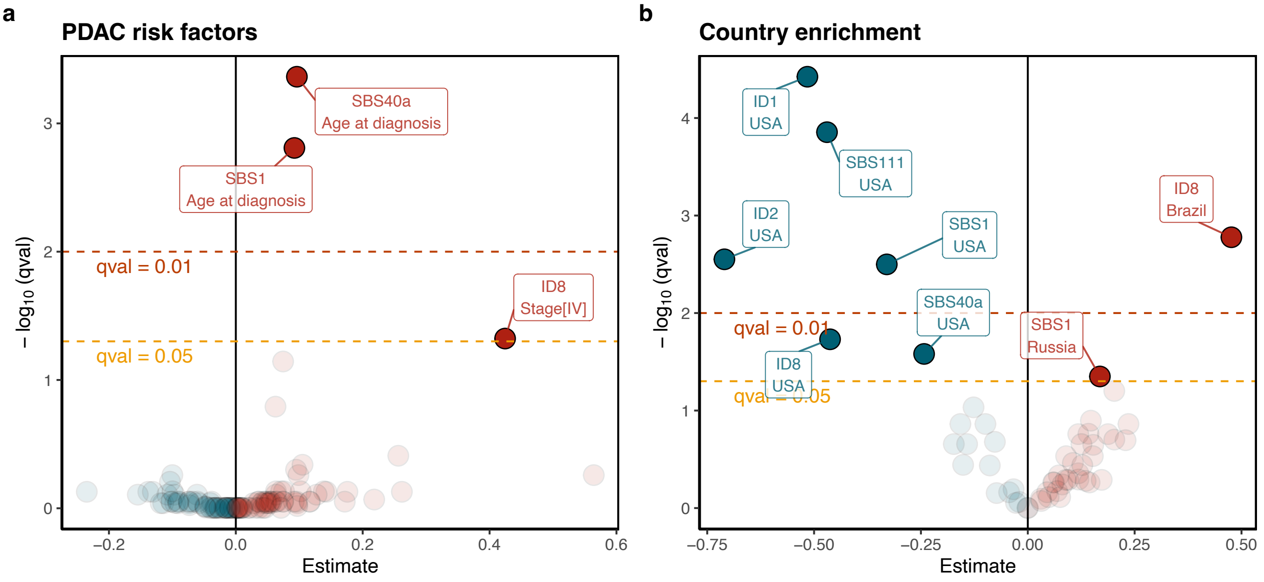

### Supplementary Fig.11. Power Calculations

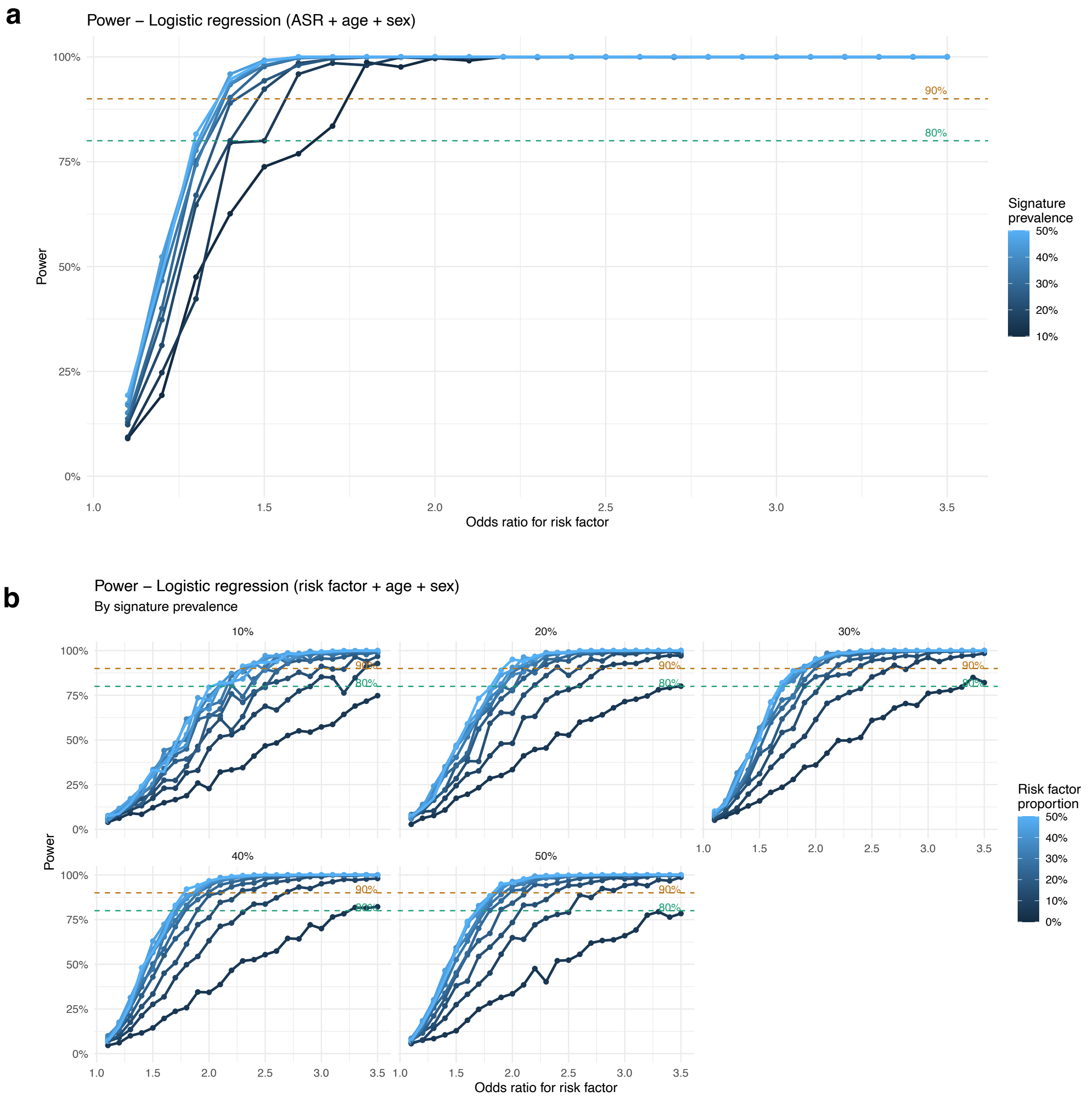
